# Detection of prodromal Parkinson’s disease using a urine proteomics panel and machine learning

**DOI:** 10.1101/2023.09.14.23295447

**Authors:** Jenny Hällqvist, Sebastian R. Schreglmann, Kristina Kulcsarova, Matej Skorvanek, Eva Feketeova, Mie Rizig, Brit Mollenhauer, Paola Turano, Maria Giulia Bacalini, Claudio Francheschi, Nicholas Wood, Kailash P. Bhatia, Kevin Mills

## Abstract

Parkinson’s disease is a progressive neurodegenerative disorder and idiopathic REM-sleep behaviour disorder (iRBD) has been identified as its single most specific early symptom. To facilitate the screening of individuals at high risk to develop Parkinson’s disease, we developed a multiplexed panel of urine proteomics using machine learning and targeted mass spectrometry to detect iRBD. Random urine samples from clinically and genetically well characterized patients with iRBD, idiopathic and hereditary forms of Parkinson’s disease, and matching controls, collected in two academic centres, were analysed in a standardized way.

First, a biomarker discovery and exploratory comparison of samples from randomly selected idiopathic Parkinson’s patients and age/sex-matched healthy controls were proteomically profiled and quantified (> 2500 proteins). The most differentially expressed biomarkers were combined into a high-throughput, multiplexed assay using targeted proteomics designed for use of tandem mass spectrometers for potential translation into clinical practise. This was then validated on independent patient and control samples (n=184). After detecting a major influence of sex on the proteome, we focused subsequent analyses on the larger group of available male samples (*n* = 114) and report results based on iRBD (*n* = 14), idiopathic (*n* = 35) and young-onset Parkinson’s disease (*n* = 15), carriers of LRRK2 (*n* = 10), PARKIN-gene mutations (*n* = 5), and healthy control subjects (*n* = 35). After establishing excellent compatibility between the two study sites, orthogonal partial least squares discriminant analysis (OPLS-DA) excluded a relevant effect of aging, but detected significant differences between iRBD and healthy controls (ANOVA-CV *P* = 0.002), as well as the combination of iPD/iRBD and healthy controls (ANOVA-CV *P* = 0.01).

Uni- and multivariate analyses detected a shared expression pattern for the protein biomarkers UBC, NCAM1, MIEN1, SPP2, REG1B, ITIH2, BCHE and C3 between iRBD and idiopathic Parkinson’s disease. Utilizing split train/test-datasets in a multiple-regression classifier model resulted in a mean accuracy of 78% to detect iRBD, matching iRBD’s conversion rate to Parkinson’s disease. Hierarchical clustering revealed greater similarities in urine proteomic changes between iRBD and idiopathic than monogenic Parkinson’s disease. Several proteins identified correlated either with clinical severity (e.g. VCAM1, MSN, HPX), or risk for future conversion to Parkinson’s disease (VCAM1, MSN, MYO10, HSPAIL).

This demonstrates the power of machine learning and urine biomarkers to identify iRBD patients. As we develop new therapies and interventions, the ability to detect individuals at-risk of neurodegeneration in very early disease stage will be invaluable for treatment success.

## Introduction

Parkinson’s disease is a progressive neurological disorder that affects movement and can cause a wide range of symptoms. Accurate diagnosis allows healthcare professionals to develop an appropriate treatment plan tailored to the individual’s specific needs. Early detection and intervention can help manage symptoms more effectively and as we develop new therapies and interventions, the ability to detect individuals at-risk of neurodegeneration in very early disease stage will be invaluable in future clinical trials and outcomes. Idiopathic REM-sleep behaviour disorder (iRBD) is a characteristic parasomnia manifesting by dream-enacting behaviour, unpleasant dreams and loss of REM-sleep muscle atonia ^1^. Beyond its immediate relevance as a sleep disorder, iRBD importantly has been established as the most relevant non-motor prodromal marker for Parkinson’s disease, the second most frequent cause of neurodegeneration ^2^, and is hence of particular interest to the Parkinson’s disease community.

Parkinson’s disease is currently the fastest growing neurodegenerative disorder in man ^4,5^, its prevalence and incidence predominantly increasing in an age-dependent manner ^3–6^. With an ageing population across the globe, the significant and increasing socio-economic burden ^7^ is projected to become an even more pressing burden to individual and public health ^8^. Although the clinical syndrome of Parkinson’s disease is meanwhile understood to encompass a heterogeneous multitude of aetiologies ^9,10^, the common underlying pathology is defined by a progressive accumulation of alpha-Synuclein (a-Syn) ^11,12^, possibly starting in the enteric or peripheral nervous system ^13–16^. Classical Parkinson’s disease is hence preceded by an insidious prodromal phase with a sub-threshold degree of neurodegeneration ^17^ without overt motor symptoms ^18^.

Although the identification and study of genetic risk variants has dramatically improved our understanding of the underlying pathophysiology and genetic architecture of this condition ^10,19^, so far this failed to translate into earlier or more accurate diagnosis ^20^ outside highly dedicated movement disorder centres ^21^. The search for a Parkinson’s disease biomarker has been ongoing for some time ^22,23^, and while promising candidates such as neurofilament light chain (NfL) have proven non-specific ^24^, α-Syn oligomers detection via seed amplification assays appear promising both in CSF ^25,26^, as well as blood ^27^, but depend on specialised lab expertise ^24^.

Multiple clinical trials have failed ^28^ and continue to fail to establish a disease-modifying therapy despite considerable efforts ^29–31^. Along several appreciated issues, pertaining from the ideal trial design, patient and outcome measure selection, to the unequivocal proof of neuroprotective efficacy ^32^, the core problem of detecting the condition in the pre-motor phase - before the majority of damage is done – remains ^33,34^. The identification of a biomarker of pre-symptomatic Parkinson’s disease would greatly facilitate these efforts.

Meanwhile, iRBD has been recognized as the most specific single clinical prodromal marker of Parkinson’s disease to date ^35–37^. The risk of pheno-conversion to an alpha-synucleinopathy increases with the time since iRBD diagnosis, ranging from 15-35% within 5 years ^38^, to 81% within 25 years ^39^. Hence, iRBD patient cohorts have increasingly been studied for Parkinson’s disease conversion risk and are generally considered an ideal target for future disease-modifying therapy trials ^33,34,40,41^.

The urine proteome is an interesting source for non-invasive biomarker development, as it is readily available in large quantities, the majority of itś content independent of diurnal variability and proteolytic degradation, and accessible to mass spectrometer measurement ^42^. First studies have capitalised on these characteristics for biomarker detection in patients with cognitive decline ^43,44^, neural ceroid lipofuscinosis ^45^, kidney disease ^46^, cancer ^47^, dermatomyositis ^48^ but also Parkinson’s disease ^49,50^.

In this work, we utilised the power of machine learning to create a panel of biomarkers into a scalable, multiplexed, mass spectrometry-based urine proteomics assay that can differentiate iRBD patients from healthy controls. The assay was based on the combination of eight protein biomarkers, identified by an untargeted discovery phase and candidates identified in the literature, and applied as a targeted validation amongst cohorts of iRBD, idiopathic Parkinson’s disease, asymptomatic and symptomatic LRRK2 carriers, Parkin carriers and healthy controls. We demonstrate that this panel allows the identification of iRBD and has the potential to predict clinical conversion to Parkinson’s disease.

## Materials and methods

### Study Cohorts

Study participants were recruited from neurology outpatient clinics at the UCL Institute of Neurology, London, and Safarik University and University Hospital, Kosice. In London, recruitment was performed at a specialist movement disorder outpatient clinic and consisted of consecutive patients with clinically established/probable Parkinson’s disease according to current diagnostic criteria ^51^ and established gene carrier status for LRRK2 and PRKN. Young-onset Parkinson’s disease (YOPD) was defined by motor symptom onset before 45 years of age ^52^. Age- and sex-matched healthy controls without history, signs or symptoms of parkinsonism or other neurodegenerative disease were recruited amongst patient partners. Asymptomatic LRRK2 carriers were recruited from first degree blood relatives of patients who are attending the movement disorders clinic at the National Hospital for Neurology and Neurosurgery, Queen Square, UK and known to be carriers for the LRRK2 G2019S mutation. In Kosice, participants were identified from a prospective data registry and a multistage screening for iRBD (PDBIOM study), which was diagnosed based on one or two nights of video-polysomnography (PSG) ^53,54^. In addition to extensive demographic and clinical details (as outlined below), dopamine transporter scintigraphy (DaT scan), as well as calculated probability of prodromal Parkinson’s disease (pPD) based on the updated MDS criteria ^55^ as reported previously ^53^ were recorded and assessed. Healthy controls were prospectively recruited amongst deeply phenotyped PDBIOM subjects with negative PSG and normal values of pPD probability based on the updated MDS criteria ^55^.

All urine samples from participating study sites were collected according to identical sample processing protocols. Mid-stream urine convenience samples were collected during routine out-patient appointments in high density polyethylene (UCL: R53102 Rocket URIKONE specimen pack; Kosice: 455007 VACUETTE TUBES) containers irrespective of time of day, fasting or medication status. They were aliquoted and frozen at -80°C within 2 hours of collection without prior centrifugation.

Clinical assessments included patient demographics and motor scores (MDS-UPDRS part III) for all participants, and REM sleep behaviour screening questionnaire (RBD-SQ), Sniffing-sticks 16 test (SS16), Epworth sleepiness scale (ESS), Montreal Cognitive assessment (MoCA), Beck depression inventory II (BDI II), Non-motor symptoms questionnaire (NMSS) and Parkinson anxiety scale (PAS) for iRBD patients.

The study was approved by the local institutional ethics committees of participating centres in accordance with the Declaration of Helsinki (UCL: 10/H0721/87. IRAS project ID: 62859; Kosice: 2017/EK/11024, 2015/EK/900016). All participants provided written informed consent prior to study participation.

The skew of males to females was significant in the patient cohorts (3:1 male to female ratio). Furthermore, the detection of a possible influence of sex on urine proteomics (Supplementary Figure 1) combined with insufficient numbers of female samples to confirm and perform a statistical correction for this phenomenon, all analyses were therefore concentrated on the analyses of male only samples (*n* = 114; Table 1). Healthy control subjects were recruited at both study centres and age-matched to idiopathic Parkinson’s disease patients recruited in London (*P* = 0.29) as well as iRBD patients recruited in Kosice (*P* = 0.69). Additional patients recruited in London with YOPD (*n* = 15; *P* < 0.0001), symptomatic and asymptomatic LRRK2 carriers (*n* = 10; *P* = 0.008), as well as Parkin carriers (*n* = 5; *P* = 0.001) were younger than respective controls, as well as idiopathic Parkinson’s disease patients (*P* < 0.0001). Parkin carriers had longer motor disease duration than YOPD (*P* = 0.0007), LRRK2 carriers (*P* = 0.001) and idiopathic patients (*P* < 0.0001), while motor severity as assessed by the MDS-UPDRS III, and Levodopa-equivalent daily dose (LEDD) ^56^ did not differ between groups.

**Table 1.**
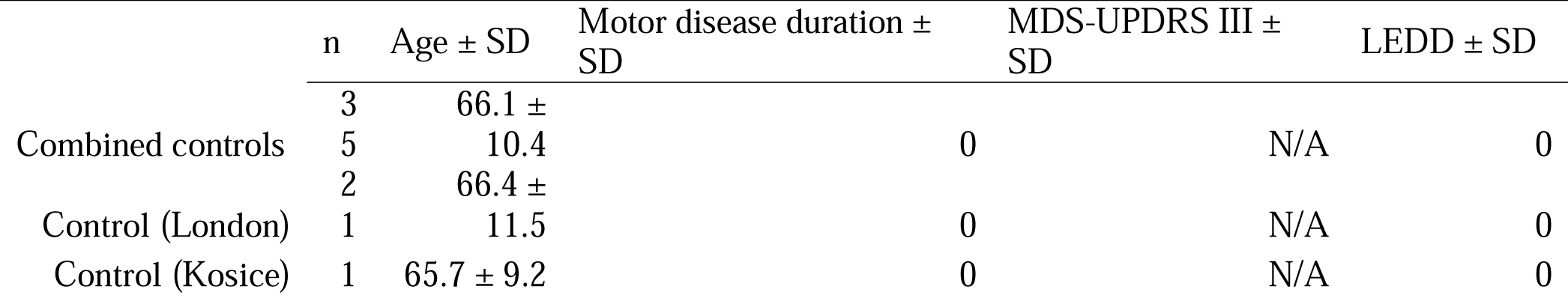
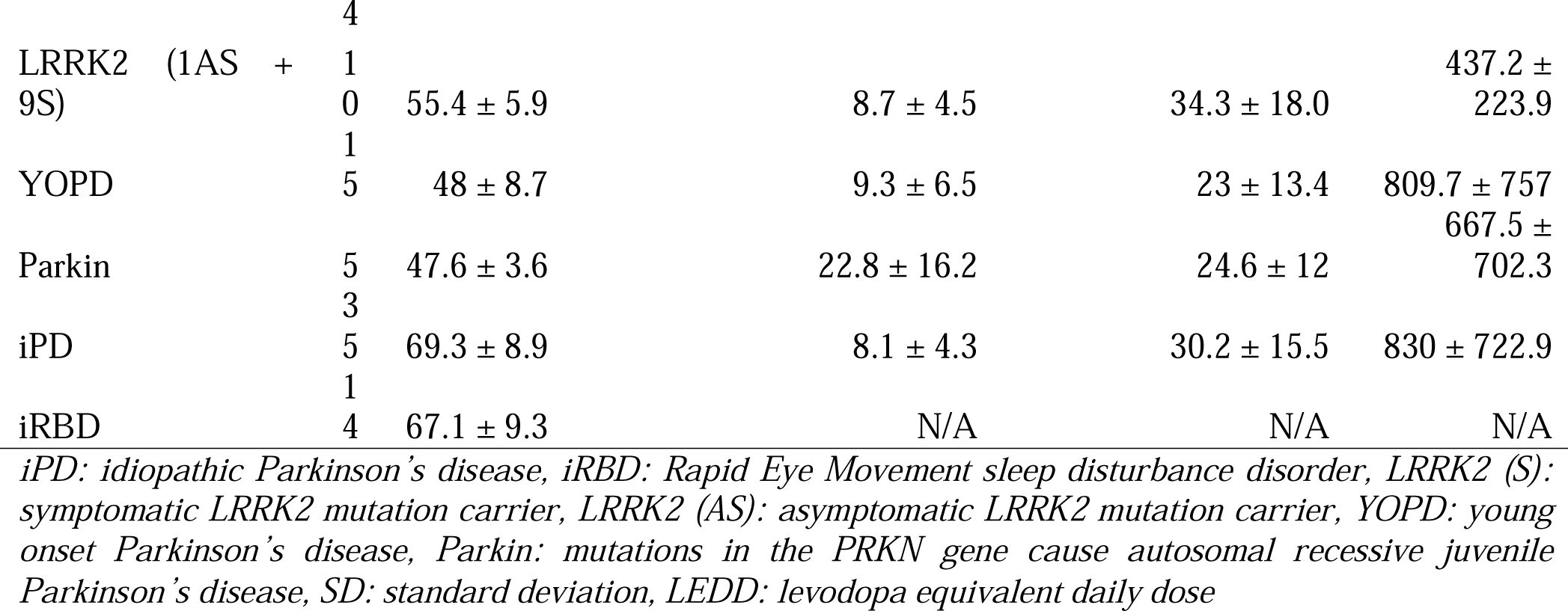
Demographic and clinical details of male only donors of urine samples analysed for targeted proteomics. The number of samples, age, motor disease duration, motor disease severity (MDS-UPDRS III score) and levodopa-equivalent dose (LEDD) ± standard deviation are reported for healthy controls, iPD, iRBD, LRRK2, YOPD and PARKIN carriers.

### Discovery Phase Cohort

The discovery cohort consisted of 9 random and well characterised patients (44% female) with idiopathic Parkinson’s disease and had been confirmed by clinical assessment and positive DaTScan (mean age 68.3 ± 7.9 years; mean motor disease duration 11.7 ± 4.1 years, MDS-UPDRS III ON medication 30.7 ± 15.1), and 10 healthy controls, matched for age (69.8 ± 10.7 years; *P* = 0.73) and sex (50% female), recruited in London.

### Targeted Proteomics Cohort

Targeted proteomic analyses were performed on samples independent of the discovery phase cohort. To detect and compare the proteomic profiles between iRBD, sporadic and two of the most frequent monogenic forms of Parkinson’s disease (LRRK2 and Parkin), as well as controls, urine samples of *n* = 184 patients from two independent cross-sectional cohorts were included in the analyses (Supplemental Table 1).

## Discovery urine proteomics

### Sample preparation

Random urine samples were thawed at room temperature, followed by vortex for five seconds. 4 mL of urine was aliquoted into 5 mL centrifuge tubes (Sarstedt, 72.701.500). The 4 mL aliquots were centrifuged at room temperature at 3760 x *g* for 30 minutes to separate the urinary sediment from solution using a Sorvall Legend RT centrifuge. 2 mL of the supernatant was transferred to Amicon Ultra-4 10 kDa molecular weight cut-off filters (Merck Millipore, UFC8010) and 2 mL Milli-Q water was added to give a final volume of 4 mL. To concentrate the urinary proteins, the samples were centrifuged at room temperature for one hour at 4445 x *g* (Sorvall Legend RT). The concentrate was transferred to a 1.5 mL centrifuge tube (Eppendorf). To ensure maximum recovery, the filters were washed with 100 μL 50 mM ammonium bicarbonate which was pooled with the concentrate. 800 μL ice-cold acetone was added to the pooled concentrate and the samples were vortexed for five seconds before overnight incubation in -20 °C. To separate the supernatant from the protein pellet, the samples were centrifuged for ten minutes at +4 °C and 16900 x *g* using a Micro-centrifuge 5424 R (Eppendorf). The supernatant was carefully pipetted off and discarded. The pellet was air dried in a fume hood for 20 minutes to evaporate residual acetone. 100 μL Milli-Q water was added to the samples and the protein pellet broken up by vigorous vortexing. The samples were thereafter freeze dried overnight, followed by tryptic digestion and solid phase extraction (SPE) as described by Bennett et al ^57^. The digested and SPE-cleaned samples were reconstituted in 50 μL 3% acetonitrile, 0.1% TFA and a peptide assay (Pierce Quantitative Colorimetric Peptide Assay from Thermo Fisher Scientific) was performed as per the manufacturer’s instructions. The peptide concentration in the samples was normalised to 1000 ng/μL before instrumental analysis by 2D-LC-MS.

### Proteomics and biomarker discovery analyses of urine from control and Parkinson’s Disease patients

Tryptic peptides were fractionated using a 2D-NanoAquity liquid chromatography system (Waters, Manchester, UK). All samples were fractionated in-line into ten fractions and analysed by label free quantitative proteomics over a 12-hour period as previously described ^58^. Fractionation was performed by reversed phase chromatography, utilising the orthogonality in peptide separation between high and low pH mobile phases. The eluted peptides were detected using a Synapt-G2-S*i* (Waters) equipped with a nano-electrospray ion source. The mass spectrometer was a Synapt-G2-Si (Waters). Data were acquired in positive MS^E^ mode from 0 to 60 minutes within the m/z range 50 - 2000. The capillary voltage was set to 3 kV and the source temperature to + 100 °C. The cone gas consisted of nitrogen with a flow of 50 L/h, the desolvation temperature was set to + 200 °C. The purge and desolvation gas consisted of nitrogen, operated at a flow rate of 600 mL/h and 600 L/h respectively. The gas in the IMS cell was helium with a flow rate of 90 mL/h. The low energy acquisition was performed applying a constant collision voltage of 4 V with a 1 second scan time. High energy acquisition was performed by applying a collision energy ramp, from 15 to 40 V, the scan time was 1 second. The lock mass consisted of 500 fmol/µL [glu1]-fibrinopeptide B, continuously infused at a flow rate 0.3 µL/min and acquired every 30 seconds. The doubly charged precursor ion, m/z 785.8426, was utilised for mass correction.

### Untargeted data processing

After acquisition, data were imported to Progenesis QI for proteomics (Waters) and the individual fractions processed before all results were merged into one experiment. The Ion Accounting workflow was utilised, with UniProt Canonical Human Proteome as a database (build 2016). The digestion enzyme was set as trypsin. Carbamidomethyl on cysteines was set as a fixed modification; deamidation of glutamine and asparagine, and oxidation of tryptophan and pyrrolidone carboxylic acid on the N-terminus were set as variable modifications. The identification tolerance was restricted to at least two fragments per peptide, three fragments per protein, and one peptide per protein. An FDR of 4% or less was accepted. The resulting identifications and intensities were exported and variables with a confidence score less than 15 and only one unique peptide were filtered out.

### Development of a multiplexed and targeted proteomic assay to validate any potential biomarkers as a test for predicting Parkinson’s Disease

The peptides included in the targeted assay were selected from the proteomic screening study as in previous work^59^. Furthermore, due to the suggested involvement of inflammation in neurodegenerative diseases, several known pro- and anti-inflammatory proteins identified from the literature were also included in the multiplexed assay (Supplementary Figure 2). This validatory test in the first instance consisted of 87 proteins, out of which a number were measured with two peptides, leading to a total of 131 unique peptides. When possible, the peptides were chosen to have an amino acid sequence length between 7 and 20. The amino acid sequences were confirmed to be unique to the proteins by using the Basic Local Alignment Search Tool (BLAST) provided by UniProt ^60^. Synthetic peptide standards were purchased from GenScript (Amsterdam, Netherlands). To establish the most optimal transitions, repeated injections of 1 pmol peptide standard onto a Waters Acquity ultra-performance liquid chromatography (UPLC) system coupled to a Waters Xevo-TQ-S triple quadrupole MS were performed. The most high-abundant precursor-to-product ion transitions and their optimal collision energies were determined manually or using Skyline ^61^. Two transitions were chosen for each peptide, one quantifier for relative concentration determination and one qualifier for identification, totally rendering 266 analyte transitions. Cone and collision energies varied depending on the optimal settings for each peptide. Each peptide was measured with a minimum of 12 points per peak and a dwell time of 10 milliseconds or more to ensure adequate data acquisition. The optimised transitions were distributed over two multiple reaction monitoring (MRM) methods, always keeping the quantifier and qualifier for each peptide in the same MRM segment.

The samples were prepared for instrumental analysis in the same way as the untargeted sample set, except for 150 ng whole protein yeast ENO1 being added prior to filtration, and that the peptide concentrations were not normalised prior to instrumental analysis. The samples were reconstituted in 50 μL 3% acetonitrile, 0.1% trifluoroacetic acid, containing 0.1 µM of isotope labelled internal standards from the following proteins (annotated by gene name): ALDOA, C3, GSTO1, RSU1, and TSP1.

### Targeted data processing

After acquisition, peak picking and integration were performed using TargetLynx (version 4.1, Waters) or an in-house application (“mrmIntegrate”) written in Python (version 3.8). mrmIntegrate is publicly available to download via the GitHub repository https://github.com/jchallqvist/mrmIntegrate. The application takes text files as input (.raw files are transformed into text files through the application “MSConvert” from ProteoWizard ^62^ and applies a LOWESS filter over 5 points of the chromatogram. The integration method to produce areas under the curve is trapezoidal integration. The application allows for retention time alignment and simultaneous integration of the same transition for all samples. Peptide peaks were identified by the blank and matrix calibration curves. The integrated peak areas were exported to Microsoft Excel where first the ratio between quantifier and qualifier peak areas were evaluated to ensure that the correct peaks had been integrated. The digestion efficiency was evaluated by monitoring the presence of baker’s yeast ENO1 in the samples, all samples without a signal were excluded from further analysis. The instrumental stability was assessed by inspecting the levels of the isotope labelled internal standard peptides. After the initial quality assessment, the urine concentrations in the samples were harmonised by probabilistic quotient normalisation (PQN) ^63^. Pooled urine quality control samples were additionally evaluated to assess the robustness of the instrumental analysis.

## Bioinformatic analysis

### Sample quality control

The untargeted and targeted datasets were inspected for outliers and instrumental drift using principal component analysis (PCA) and orthogonal projection to latent variables (OPLS) in SIMCA, version 17 (Umetrics Sartorius Stedim, Umeå, Sweden). After normalization of urine concentrations using probabilistic quotient normalization (PQN), outliers exceeding six median deviations from each variable’s median were excluded. To equalise small instrumental differences, samples within each run batch were z-scored before merging the datasets. Each peptide was thereafter filtered or averaged so that each protein was represented by a single variable. The data were evaluated for normal distribution using D’Agostino and Pearson’s method from SciPy (version 1.11.0). The dataset was inspected for outliers and instrumental drift using principal component analysis (PCA) and orthogonal projection to latent variables (OPLS) in SIMCA, version 17 (Umetrics Sartorius Stedim, Umeå, Sweden).

### Statistical analysis, visualisation and modelling

Most of the statistical analyses were performed in Python (version 3.8.16). The untargeted dataset was analysed for differences between the groups by applying Student’s two-tailed t-test. Due to the limited sample numbers, the significance threshold was set nominally to *P* < 0.05. The targeted data were evaluated for normal distribution using D’Angostino and Pearson’s method from SciPy. Significance testing between the independent groups was performed by Student’s two-tailed t-test for normally distributed variables and by Mann-Whitney’s non-parametric U-test, both from SciPy’s stats package, for non-normally distributed variables. The Benjamini-Hochberg multiple testing correction false discovery rate (FDR) procedure (Statsmodels version 0.14.0) was applied with alpha set to 0.1. Fold-changes were calculated by dividing the means or medians of the affected groups by the control group. Correlation analyses in the targeted data were performed by Spearman’s correlation (SciPy). Box plots and hierarchical clusters were produced using the Seaborn (version 0.12.2) and Matplotlib (version 3.7.0) libraries. All multivariate analyses were performed in SIMCA version 17 (Sartorius Umetrics). OPLS and OPLS-discriminant analysis (OPLS-DA) models were evaluated for significance by ANOVA p-values and by permutation tests applying 1000 permutations, where p < 0.05 and p < 0.001 were deemed significant, respectively. Data were analysed for pathway enrichment and Gene Ontology (GO) annotations using DAVID Bioinformatics Resources (2021 build). Networks were built in Cytoscape (version 3.8.0) applying the “Organic layout” from yFiles. Logistic regression (scikit-learn, version 1.2.2, ^64^) was utilised for multiple-regression classifier modelling. The data was split into two 50% parts, one for model training and one for prediction, using the function train-test-split from Scikit-learn. Cross-validation was performed using a stratified k-fold with five splits and the cross validated scores for the model were calculated as mean and standard deviation. ROC curves were constructed from each predictor and from the combined predictors. Plots of the data were constructed using the Matplotlib package (version 3.6.0) ^65^.

## Data availability

Chromatograms and raw data files from the targeted proteomic analysis are available via the public Panorama repository https://panoramaweb.org/urine_irbd_trgt_proteomics.url. Further data are available from the corresponding author by reasonable request.

## Results

### Proteomic discovery phase and development of a targeted assay

Label free quantitative proteomics was performed using two-dimensional in-line nano-liquid chromatography in ten fractions using QTOF MS^E^ (Waters, UK). This deep phenotyping proteomic analysis of the nine idiopathic PD (iPD) and 10 Control urine samples identified > 2500 proteins, of which 89 were differentially expressed between groups on a significance level of 95% (Figure 1A). Pathway analysis of the differentially expressed proteins demonstrated that several inflammatory pathways were enriched, along with pathways relating to mitochondrial dysfunction and Parkinson’s disease (Figure 1B). Based on the discovery phase findings, we constructed a targeted mass spectrometric assay in which the included proteins were selected by a mixture of greatest fold-change, p-value significance and top candidates discovered by other research groups identified via literature studies ^59^. This resulted in a panel of 87 proteins (Figure 1C).

**Figure 1.**
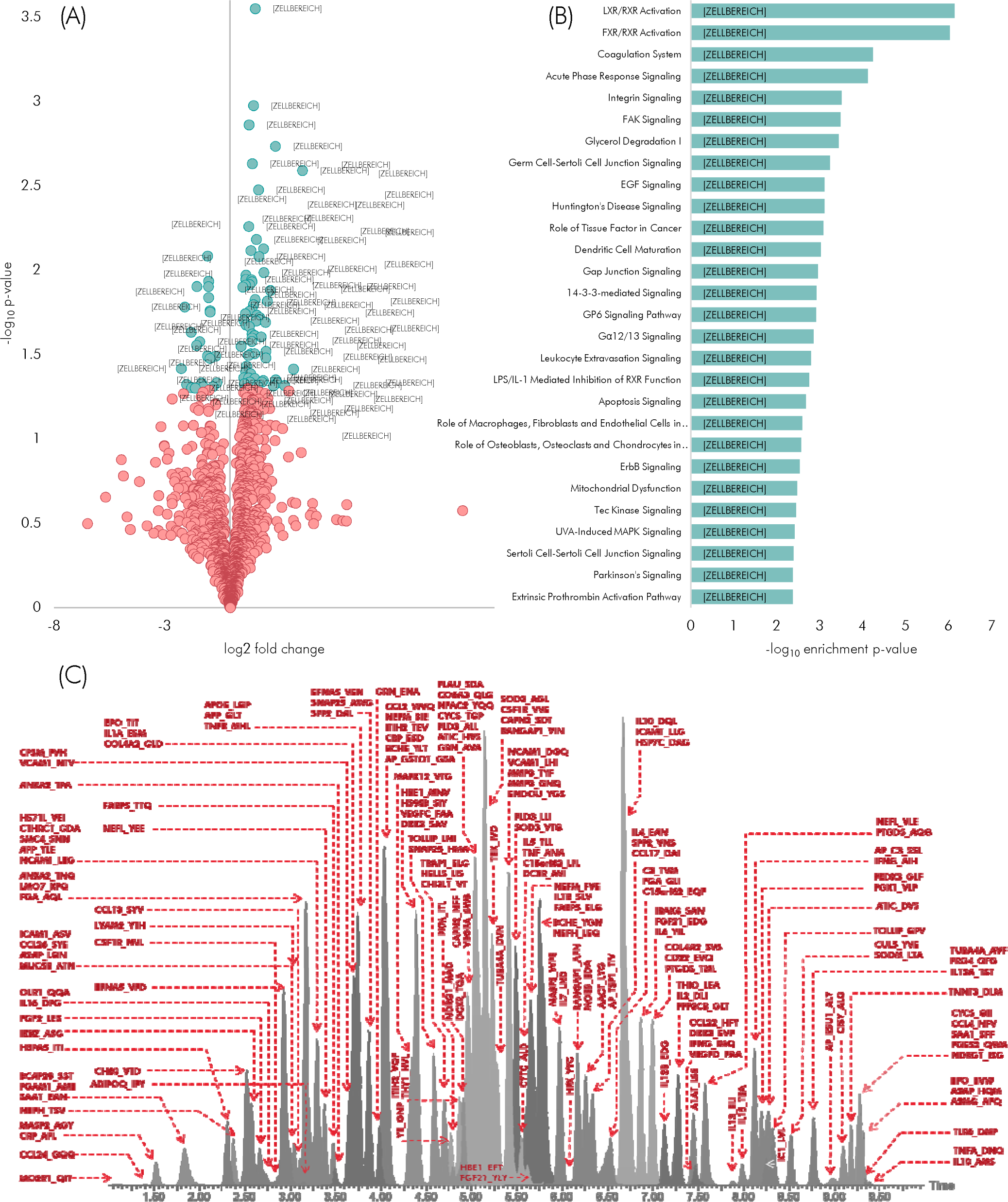
Discovery phase analysis and targeted proteomic panel development. **(A)** Volcano plot depicting 89 proteins differentially expressed between Parkinson’s patients and controls in the discovery phase. (**B**) Significantly enriched pathways based on the findings from the untargeted proteomic analysis. **(C)** Based on proteins identified through further screening studies, the targeted protein panel was developed, consisting of 87 poteins.

### Targeted proteomics

A targeted and scheduled proteomic assay was developed using ultra-high performance liquid chromatography (UPLC) system coupled to a triple quadrupole mass spectrometer operating in multiple reaction monitoring mode (MRM). The targeted assay was applied to a new cohort of urine samples consisting of controls, LRRK2 mutation carriers, YOPD, Parkin mutation carriers, idiopathic PD and iRBD. In total, we could reliably detect and quantitate 50 proteins in each of the urine samples. Quality control samples, consisting of pooled control and patient samples, demonstrated a coefficient of variation ranging from 3% to 28%, with 94% of the measured proteins attributed with a coefficient of variation of 15% or less (Supplementary Fig. 3).

### Control sample compatibility and influence of age

To determine potential inter-centre variation, age-matched healthy control subjects from the two independent participating centres were compared (London: *n* = 49; Kosice: *n* = 17, 64.4 ± 10.6 years vs. 65.5 ± 9.1 years; *P* = 0.7). We compared the multivariate expression in the control group by orthogonal projection to latent structures discriminant analysis (OPLS-DA) with centre as the discriminant variable. The model was non-significant and demonstrated that no discernible multivariate expression difference could be identified between the centres (*P* = 1.0). In addition, univariate analysis applying Student’s two-tailed t-test, followed by the Benjamini-Hochberg procedure for multiple testing correction (FDR = 10%), showed that none of the proteins demonstrated a significant difference between the two centres. Taken together, the inability to discriminate between the control samples both multi- and univariately demonstrates excellent compatibility between the centres. We next explored if there was an effect of age on protein expression through an OPLS model, where age was set as y. This model was non-significant with ANOVA-CV and permutations *P* > 0.05, also demonstrated that no age-related covariation between the variables could be found.

### Proteomic identification of idiopathic REM-sleep behaviour disorder

Principle Component Analyses (PCA) was used for an initial overview of the targeted proteomic data and demonstrated that no patient sample clusters were readily identifiable (Figure 2). Investigating the discrimination between iRBD samples and healthy controls, an OPLS-DA model of these two sample groups was constructed. After visual inspection of the cross validated (CV) scores and permutations evaluation (*P* < 0.001), the model was found significant (ANOVA-CV *P* = 0.002). The proteins with strongest influence on the discrimination between the groups were UBC, VCAM1, MIEN1, GOLM1 and ICAM1 – upregulated in iRBD, and ITIH2, APOE, BCHE, CCL4 and PLD3 – downregulated in iRBD (Figure 3B). We further utilised the discriminating OPLS-DA model of iRBD *versus* control to evaluate the level of shared protein expression, compared to control, with the other Parkinsonian sample groups by prediction testing. This resulted in rather low proportions of the groups predicted as iRBD (57% for iPD; 60% for YOPD and LRRK2; 80% for Parkin), thus implying a limited degree of shared expression between the proteins typical for iRBD and Parkinsonian disorders in this sample set.

**Figure 2.**
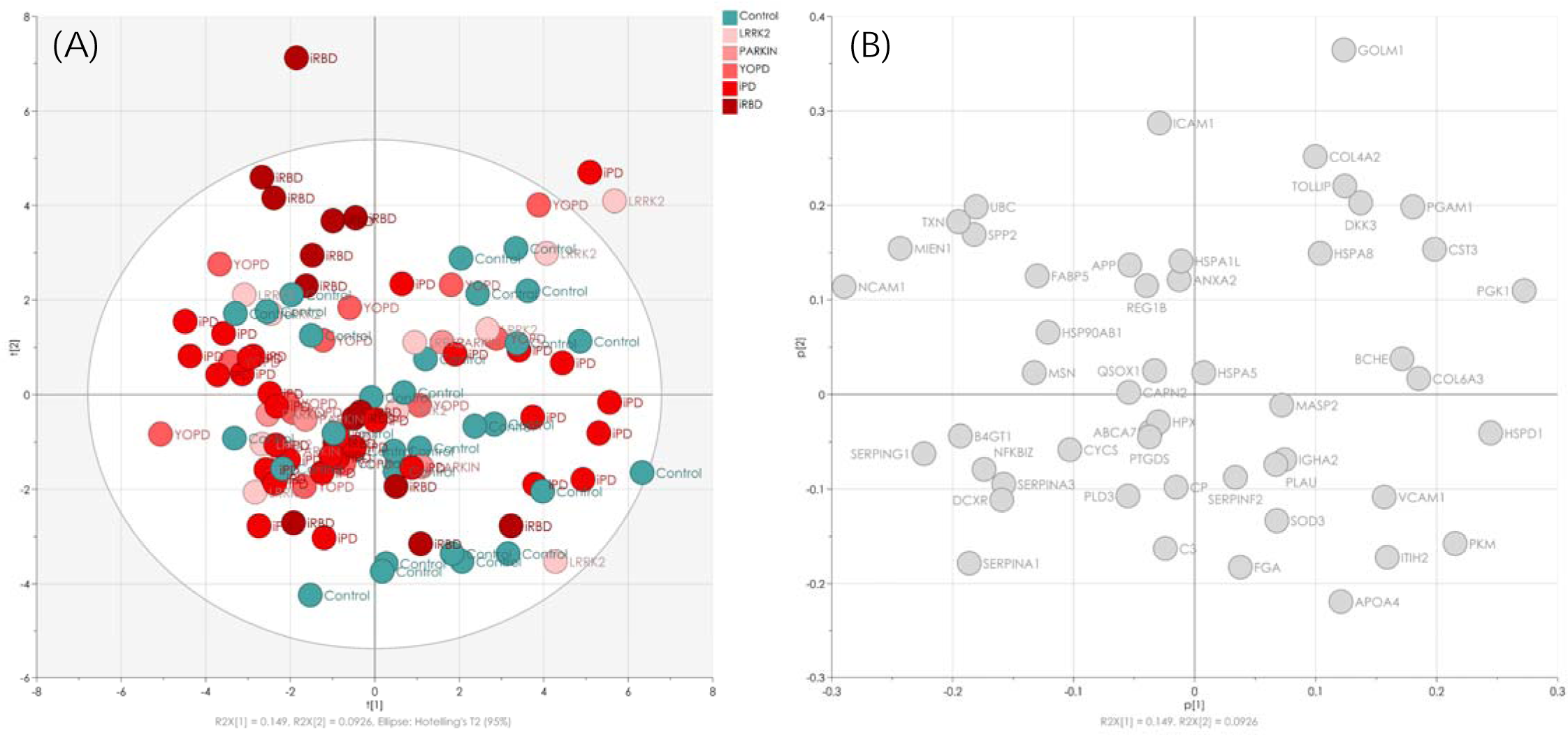
Targeted urine proteomics of Parkinsonian samples. **(A)** A principal component analysis (PCA) of targeted urine proteomics in the Parkinsonian sample groups iPD, YOPD, Parkin and LRRK2, a prodromal group consisting of iRBD, and healthy controls. The scores demonstrated no clear differences between the sample groups, indicating subtle differences in protein expression between the patient groups and controls. **(B)** PCA loadings showing the proteins responsible for the samples’ scores distribution.

**Figure 3.**
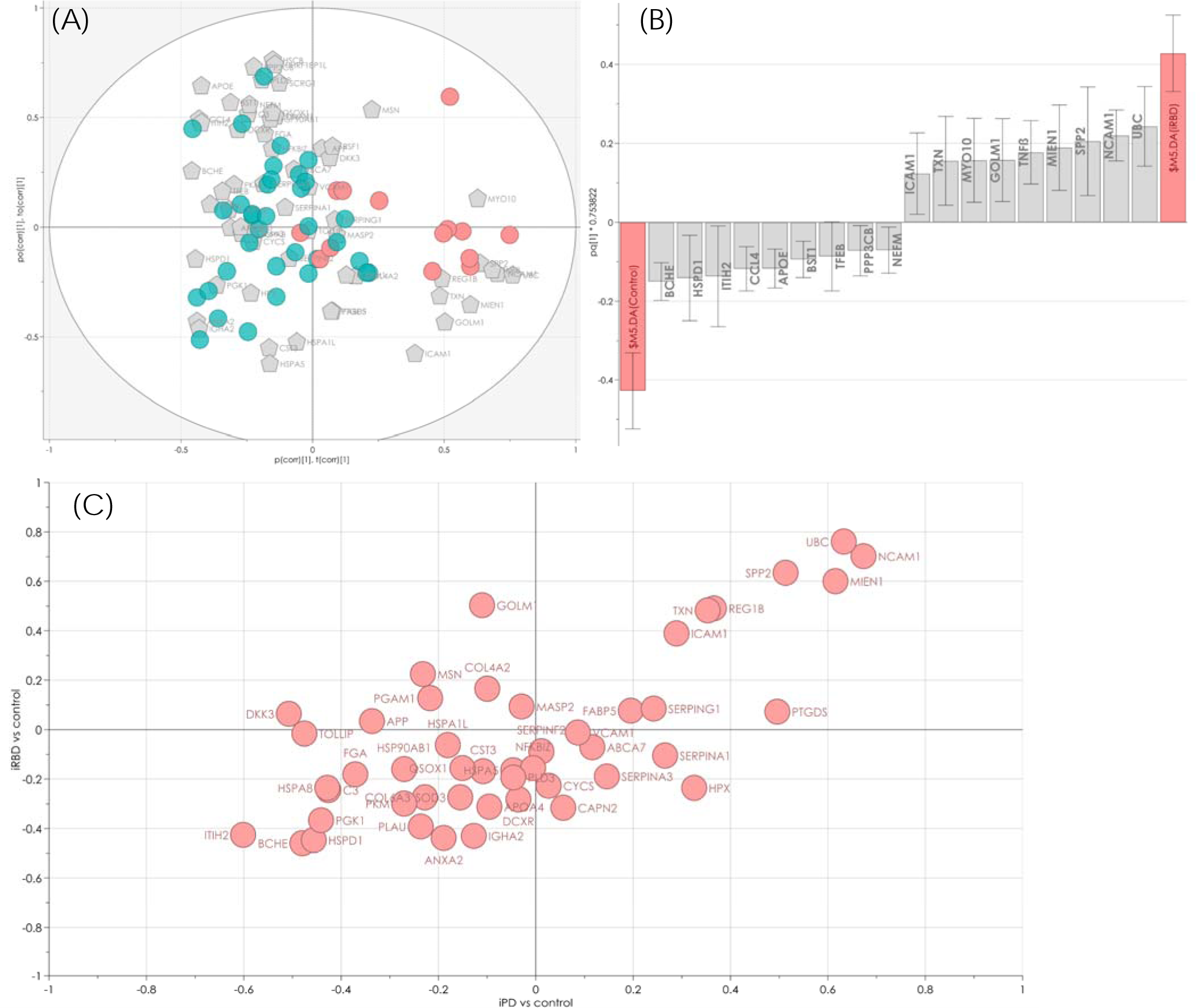
Orthogonal partial least squares discriminant analysis results of targeted proteomic analysis in iRBD and Parkinson’s disease. Displayed results represent orthogonal partial least squares discriminant analysis (OPLS-DA) comparisons of idiopathic REM-sleep behaviour disorder (iRBD) patients versus controls, and their communalities with idiopathic PD (iPD) versus controls. **(A)** The iRBD OPLS-DA model discriminates significantly against controls (ANOVA-CV *P* = 0.002), with differences visualised in a iBiplot (a second orthogonal component has been added for visualisation purposes). **(B)** Predictive loading plot showing the proteins with strongest effect on the discriminating ability of the model. **(C)** Shared-and-unique structures plot of the two models (i) iRBD versus control, (ii) and iPD versus control, demonstrated that the two models share characteristics with, among others, UBC, NCAM1, MIEN1, SPP2 and REG1B being positively correlated with the disease groups, and ITIH2, BCHE and C3 negatively correlated.

We similarly constructed an OPLS-DA model comparing individuals with iPD *versus* healthy controls, which proved no significant differences (*P* = 0.099). However, comparing iPD + iRBD as one group versus healthy controls showed weakly significant differences (*P* = 0.01), whereas an OPLS-DA comparison between iPD *versus* iRBD could not differentiate between the two groups (*P* = 1). This indicates that although the urine proteomic expression profile was not distinct enough to differentiate iPD individually from healthy controls, there is a relevant overlap in urine protein expression between iPD and iRBD. Assessing the shared similarities between these groups and the differences to healthy controls, a shared-and-unique-structures plot (Figure 3C) of the models (i) iRBD *versus* control, and (ii) iPD *versus* control, was created. This showed that despite differences between iPD and iRBD, they indeed shared similarities in comparison to healthy controls – with, among others, UBC, NCAM1, MIEN1, SPP2 and REG1B being positively correlated with the disease groups, and ITIH2, BCHE and C3 negatively correlated.

Finally, logistic regression was used to build a multiple-regression classifier model to differentiate between iRBD and healthy control samples. To achieve this, we utilised 50% of the samples for model training and predicted the remaining samples. This resulted in a classification accuracy of 80% using ten of the most significant variables from the comparison between iRDB and controls (UBC, BCHE, ITIH2, NCAM1, REG1B, PKM, ANXA2, SPP2, MIEN1, and ICAM1). Cross-validation of the model using five different splits of the data resulted in a mean accuracy of 78% ± 7.4%. A receiver operating characteristics (ROC) curve was then constructed and demonstrated that the combined panel of proteomic predictors resulted in greater classification accuracy than each predictor alone (Figure 4). To assess the similarities between the comparators in this iRBD model and Parkinsonian samples, we evaluated how the clinically diagnosed Parkinsonian groups were predicted in the logistic regression model. Prediction as belonging to the iRBD class was again generally low in all groups (YOPD 20%; LRRK2 30%; Parkin 20%, iPD 23%). Similar to before, this suggests that there is limited shared protein expression between these groups.

**Figure 4.**
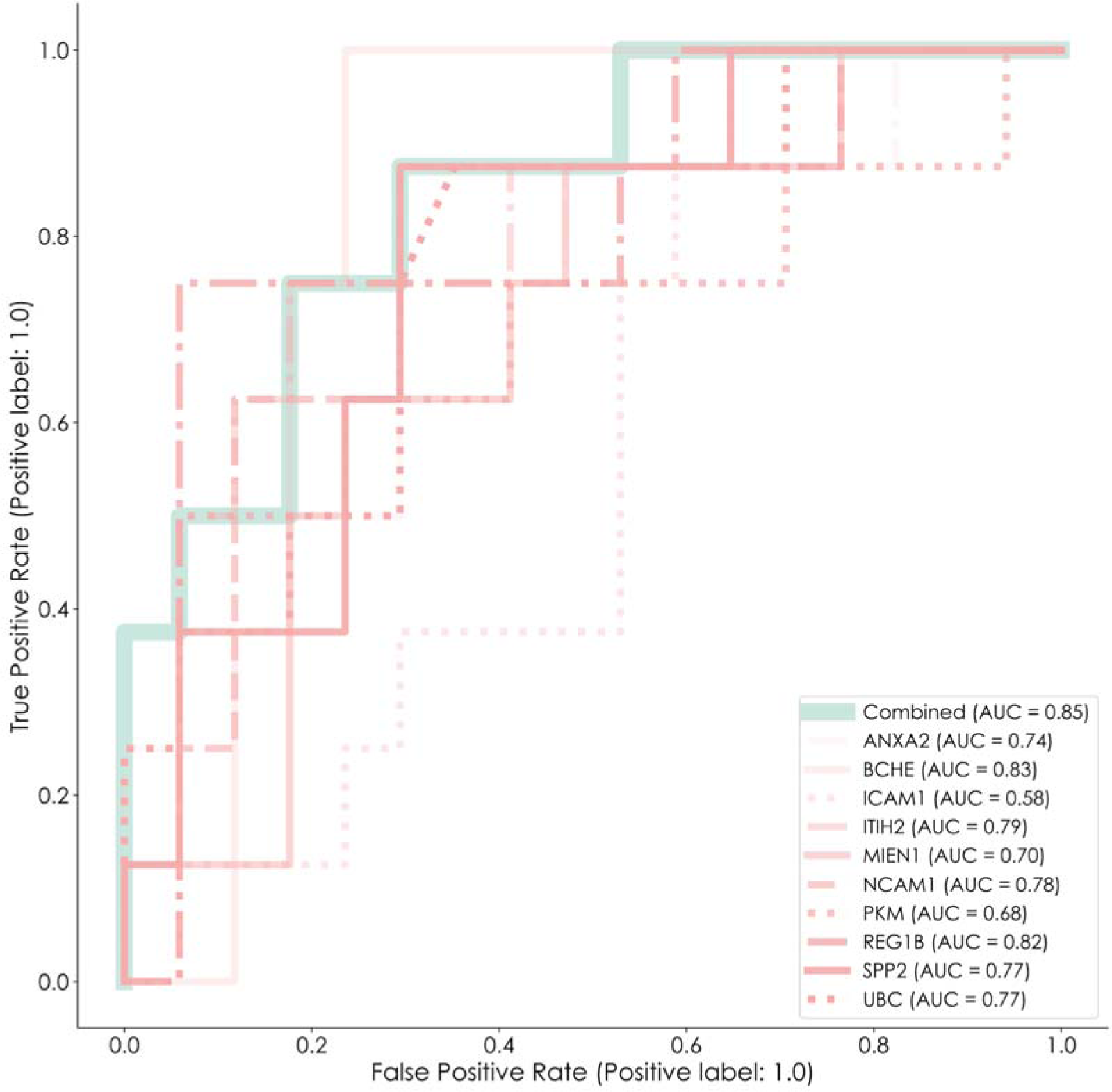
Receiver operating characteristics curve of the targeted urine proteomic panel to differentiate iRBD from healthy controls. ROC plot showing the results from a logistic regression classifier, using the individually significant proteins separately, and in combination.

### Univariate proteomic differences and clustering

The analyses of univariate protein expression differences between groups demonstrated that the iRBD group, representing prodromal PD, showed the largest differences compared to healthy controls. In total, 14 proteins were differentially expressed in the iRBD group, seven in iPD, four in LRRK2, three in Parkin and six in YOPD (Supplementary Figure 4). When assessed together, seven out of 50 measured proteins were differentially expressed in at least two of the sample groups when compared to healthy controls: ANXA2, BCHE, ITIH2, PKM, REG1B, SPP2 and UBC. All seven proteins were identified in the iRBD group, and all showed a reduced expression compared to controls (Supplementary Figure 5).

In order to explore similarities and differences on a proteomic level, a hierarchical cluster analysis was performed using the signed p-values resulting from the univariate comparisons of the disease groups (Figure 5). The resulting clustering identified two top-level clusters which grouped the proteins by direction of expression change (higher versus lower). Consistent with the above OPLS-DA analyses, the largest similarities in the overall proteomic expression profile were found between iRBD and iPD sample groups. According to this, the proteomic profile in Parkin carriers was distinct, with YOPD and LRRK2 samples clustering in between.

**Figure 5.**
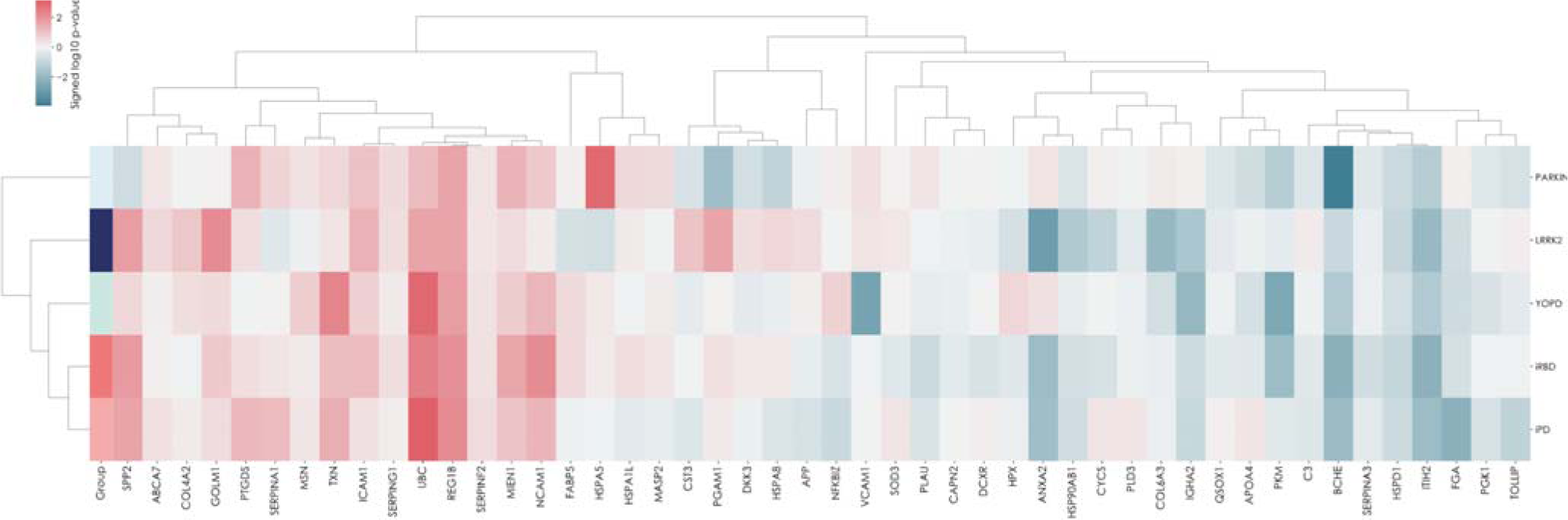
Hierarchical clustering of the urine proteome by diagnosis/genotype. Hierarchical clustering of the signed log10 p-values from the comparisons of respective parkinsonian groups and iRBD to control. The clustering method was set to “weighted” and the metric was “cosine.” The row-wise clustering of the results from the sample groups demonstrates that iPD and iRBD share characteristics, also to a lesser degree with YOPD and LRRK2. PARKIN does not make part of this general cluster. Column-wise, the proteins are separated into two top-level clusters, generally characterised by higher values in the Parkinsonian conditions compared to control to the left and lower values to the right.

### Clinical correlates of proteomic changes

To aid the interpretation of whether changes in the proteome are clinically meaningful, we explored any potential correlation with clinical metrics in the respective patient groups. In the Parkinsonian patient groups (iPD, YOPD and Parkin), correlations were evaluated between demographics, clinical metrics, and protein expression (Table). Unsurprisingly, motor disease severity (MDS-UPDRS III) correlated positively with age and motor disease duration, as well as levodopa equivalent daily dose (LEDD). Similar positive correlations were established for age with LEDD, Motor disease duration with MDS-UPDRS III & LEDD, and MDS-UPDRS III score with LEDD.

Relating proteomic changes with clinical metrics (Table 2), signals with at least two indicators of PD severity were identified for VCAM1, MSN and HPX. VCAM1 was found to positively correlate with MDS-UPDRS III and motor disease duration, MSN negatively correlated with motor disease duration and LEDD, HPX negatively correlated with MDS-UPDRS III and LEDD. Furthermore, MDS-UPDRS III negatively correlated with ICAM1. Motor disease duration was found positively correlated with ITIH2, and negatively with SERPING1, NCAM1, and TXN. Age was positively correlated with APOA4, BCHE and IGHA2, and negatively correlated with MASP2 and REG1B. Several of the identified proteins in the Parkinsonian samples were found to correlate with each other (Supplementary Figure 6).

**Table 2.** Clinical and proteomic correlations in Parkinsonian patients. Correlations with clinical MDS-UPDRS III score, motor disease duration and LEDD. The correlation was evaluated in the pooled group iPD, YOPD, LRRK2 and PARKIN. Spearman correlation was utilised and no post-hoc correction of the *P*-values was performed. The arrows indicate if the correlation was positive or negative.

|  | Age | Motor disease duration | MDS-UPDRS III | LEDD |
| --- | --- | --- | --- | --- |
| Age |  |  | 0.02 (↑) | 0.03 (↑) |
| Motor disease duration |  |  | 0.01 (↑) | 0.0006 (↑) |
| MDS-UPDRS III (ON) | 0.02 (↑) | 0.01 (↑) |  | 0.02 (↑) |
| LEDD | 0.03 (↑) | 0.0006 (↑) | 0.02 (↑) |  |
| APOA4 | 0.007 (↑) |  |  |  |
| BCHE | 0.002 (↑) |  |  |  |
| HPX |  |  | 0.05 (↓) | 0.006 (↓) |
| ICAM1 |  |  | 0.01 (↓) |  |
| IGHA2 | 0.009 (↑) |  |  |  |
| ITIH2 |  | 0.005 (↑) |  |  |
| MASP2 | 0.02 (↓) |  |  |  |
| MSN |  | 0.02 (↓) |  | 0.04 (↓) |
| NCAM1 |  | 0.02 (↓) |  |  |
| REG1B | 0.02 (↓) |  |  |  |
| SERPING1 |  | 0.03 (↓) |  |  |
| TXN |  | 0.02 (↓) |  |  |
| VCAM1 |  | 0.02 (↑) | 0.05 (↑) |  |

Among iRBD patients, we in particular evaluated clinical metrics representing the risk for future conversion to PD (Figure 6A). In this regard, we found that dopamine transporter (DaT) scan results negatively correlated with DKK3, TNFß, MYO10, HSPA1L, MSN, and positively correlated with VCAM1. The total likelihood ratios (LR total), as calculated based on the updated MDS research criteria for pPD, were negatively correlated with MSN, C3, SPP2, MYO10, HSPA1L and MSN, and positively correlated with PLAU, CST3, HPX and VCAM1. The calculated probability to develop PD (pPD; %) was negatively correlated with SPP2, GOLM1, MYO10, HSPA1L, C3, UBC, PGAM1, and PPP3CB, and positively correlated with IGHA2, CYCS, APOA4, PLAU, CST3, HPX and VCAM1. Across these analyses, four proteins were found to be overlapping in their correlation with risk for future conversion to PD, namely VCAM1 (positive) as well as MYO10, HSPA1L and MSN (negative; Figure 6B).

**Figure 6.**
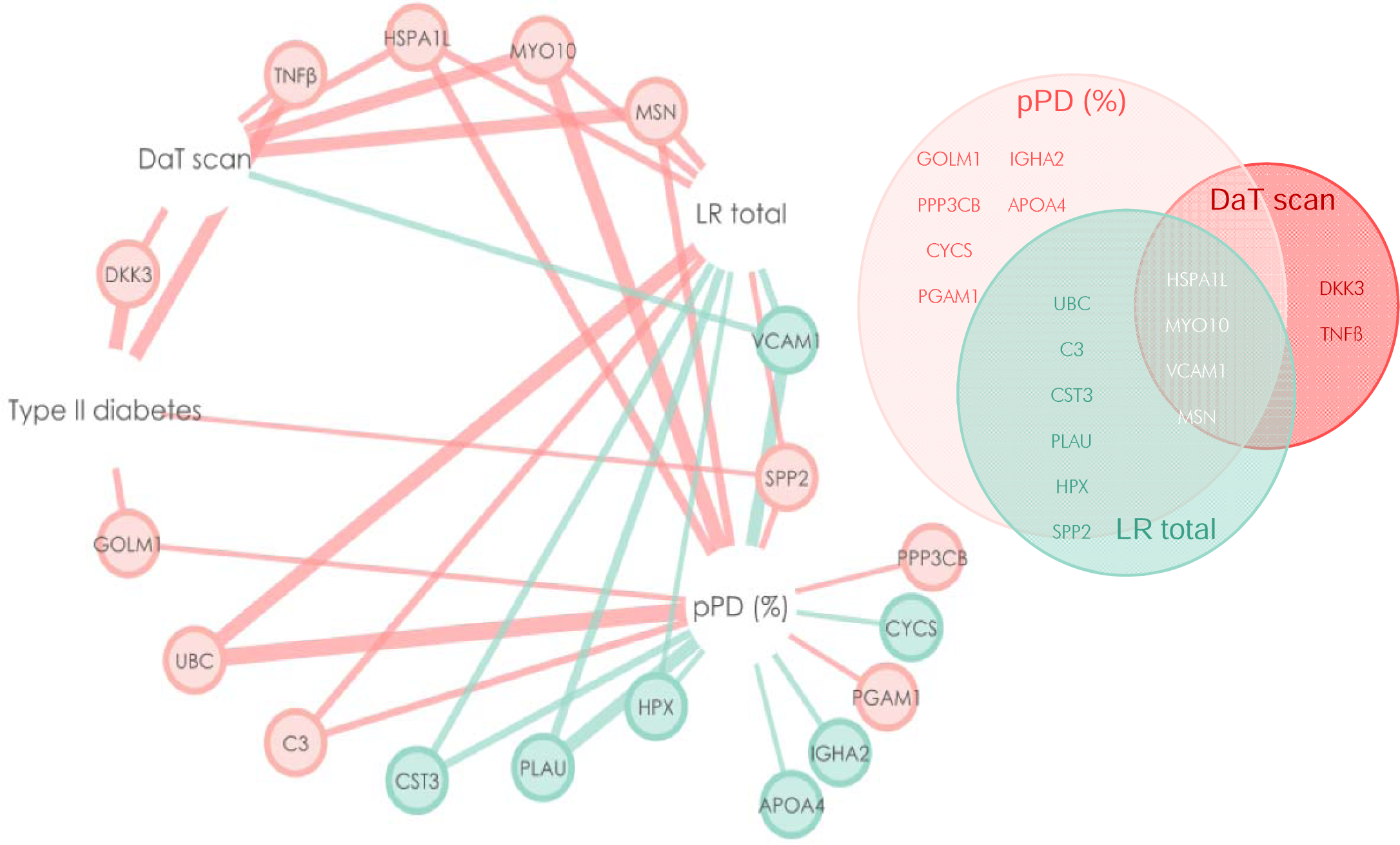
Clinical correlations with proteomic changes in iRBD samples. (**A**) Network representation of significant correlations with clinical measures for iRBD and proteins measured by targeted proteomics. Pink lines represent negative correlation and green lines positive correlations. Line thickness represents -log10 correlation p-value. (**B**) Euler diagram showing the overlapping correlations of the proteins significantly correlated with the clinical measures. HSPA1L, MYO10, MSN and VCAM1 were correlated with all three clinical measures. (Spearman correlation, no post-hoc p-value correction)

## Discussion

### Development of proteomic panel for iRBD identification

In this work we present a panel of urine proteomic markers that is able to detect iRBD but also shows overlap with similar proteomic changes in idiopathic and hereditary forms of PD. This demonstrates the power of using a combination or panels of biomarkers with machine learning. By analysing multiple biomarkers simultaneously, machine learning models can uncover intricate patterns that may not be evident to human observers. These models can learn the associations between biomarker profiles and disease states, allowing for more accurate diagnosis and prognosis. Our two-step biomarker panel development approach, capitalizes on the analytical power of untargeted proteomics followed by a validatory assay performed on triple quadrupole mass spectrometers. This strategy has the added advantage of potential for the rapid translation of these biomarkers into clinical laboratories, as most large centres have this mass spectrometry platform available.

Based on the full proteomic profiling of a small exploratory PD cohort in combination with protein targets of inflammation and neurodegeneration identified from the literature, the unbiased urine proteomic panel developed in this work overcomes the limited scope in targeted biomarker approaches based on a-priori assumptions ^22,23,25,26^. Our exploratory untargeted analysis resulted in a much deeper proteome coverage (>2500 proteins), quantifying roughly twice as many peptides as previous studies ^49,66^. Our two-step approach follows previous own work in the field of neurometabolic diseases ^45^, and Parkinson’s disease, where we recently developed a multiplexed, mass spectrometry-based plasma proteomics test essay to differentiate de-novo Parkinson’s disease patients from healthy controls with excellent classification accuracy (ref BioRxiv?). The current work expands this to urine proteomics and pre-symptomatic testing, and serves as proof-of-concept for a two-step approach for proteomic biomarker development.

In its ideal form, a biomarker is easily accessible, stable, correlates with disease severity and is scalable for a standardized adaptation across institutions. As a potential biomarker source, our laboratory has demonstrated that urine has several conceptional advantages, along ease of sampling ^42^ for the use in targeted proteomic analyses and the stratification of disease progression ^67^. In particular, due to the high salt/urea concentration, the urine proteome is relatively stable in comparison to blood-based assays regarding circadian variability and influence of food intake ^68^. It is easy to ship requiring no precautions and is easily obtainable in large amounts and, in contrast to plasma and serum, urine requires no phlebotomy or sample preparation such as centrifugation.

Our results demonstrate a certain overlap in the urine proteomic changes in iRBD and Parkinson’s disease with respect to healthy controls. This is in line with the well documented epidemiological association between the two clinical diagnoses, with over 80% of iRBD patients going to develop Parkinson’s disease ^1,37,69,70^. In particular, we detected that between iPD and iRBD, UBC, NCAM1, MIEN1, SPP2, REG1B, TXN, ICAM1, PTGDS, FABP5 and SERPING1 were positively, and ITIH2, BCHE and C3 negatively correlated with disease. From the above, NCAM1, ITIH2, and BCHE were also found to correlate with clinical markers of disease severity in Parkinson’s disease, strengthening a likely mechanistic link: NCAM1, involved in cell adhesion, axon development and neuron projection, has been previously found to be downregulated in plasma of drug-naïve Parkinson’s disease patients ^71^. ITIH2, a plasma serine protease inhibitor involved in extracellular matrix stabilization, has been found upregulated in the substantia nigra in a 1-Methyl-4-phenyl-1,2,3,6-tetrahydropyridin (MPTP) mouse model of PD ^72^. BCHE is a known bioscavenger for pesticides - its serum activity has previously been shown to be a moderate biomarker for Parkinson’s disease ^73^, while the K-variant BCHE polymorphism increases disease risk in pesticide-exposed individuals via reduced serum activity levels ^74^.

Overlapping with the above, in the iRBD samples we were able to detect a correlation between at least one measure for future conversion to Parkinson’s disease with UBC, SPP2, and C3. None of these proteins have previously been implicated with iRBD pathophysiology, and for SPP2 the protein function is unknown so far. UBC is of major importance for protein homeostasis through its role in the ubiquitin-proteasome system, which is impeded early on in the Parkinson’s disease process ^75^. C3 plays a central role in the activation of the microglia-complement-phagosome pathway, and has been reported to be elevated in the CSF ^76^, as well as reduced in plasma in Parkinson’s disease ^77^.

This further strengthens a likely mechanistic link between these proteins and the underlying developing a-Syn pathology. Mere correlation between a clinical outcome and a biomarker does not constitute a valid surrogate ^78^. However, the fact that in addition to the above-described proteomic convergence between diseases we detected half of the above listed proteins (ICAM1, PTGDS, C3 and SERPING1) similarly in plasma of drug-naïve Parkinson’s disease patients (ref Brit’s paper), serves as external validation of the identified targets and their likely pathophysiological relevance. Importantly, these proteins have also been implicated in the Parkinson’s disease process, and in particular related neuroinflammation – ICAM1 as a prominent marker of neuroinflammation, involved in sustained substantia nigra inflammation ^79^ and T-cell infiltration ^80^. ICAM1 has been found elevated in CSF in Parkinson’s ^81^, but also Alzheimer’s disease ^82^ and thus might not be specific to a-Syn pathology. Prostaglandin 2, the product of PTGDS, has been implied in the circadian system and regulation of the sleep/wake cycle ^83,84^, offering a pathophysiological link to sleep-related symptoms. Interestingly, PTGDS has also been identified to correlate with abnormal immune cell composition in Parkinson’s disease ^85^ and regulate the anti-inflammatory function of astrocytes, a mechanism involved in DJ-1 related hereditary Parkinson’s ^86^. SERPING1 has similarly been described to regulate the loss of dopaminergic neurons by regulating astrocyte expression levels in the substantia nigra of MPTP-treated mice ^87^.

Overall, the majority of marker proteins identified between iRBD and Parkinson’s disease hence point at a major involvement of the immune system – well established and reviewed in ^88^ - signs of which have recently been also detected in iRBD patients’ peripheral blood immune cell composition ^89^ and substantia nigra immune activation ^90^. The role of the immune system in iRBD, but also non-manifesting carriers of Parkinson’s disease causing mutations is increasingly recognized ^91^, and our findings are in line with this.

Our results establish urine proteomic differences between idiopathic, young-onset and different hereditary forms of Parkinson’s disease, as previously described for LRRK2 in CSF samples ^66^. Interestingly, although LRRK2 and PARKIN pathophysiology is overlapping in mitochondrial dysfunction, the proteomic profile for PARKIN was distinct and clustered separately from all other entities. The limited number of included monogenic cases warrants cautious interpretation, and greater sample numbers are needed for definitive conclusions.

### The ability of the test to detect individuals who are at risk of developing Parkinson’s Disease

Research into biomarkers in the field of Parkinson’s follows diagnostic (detecting established disease), monitoring (detecting change in severity) and prognostic (prediction of future disease development) aims. The ‘shutting the stable door after the horse has bolted’ hypothesis has gained much traction in neurodegenerative disease research. As it is appreciated that the majority of damage through neurodegeneration has already occurred once motor symptoms are present ^33^, and interventions hence less likely to salvage neuronal function, the pre-motor phase of the disease has become the focus for biomarker development ^22,37^. Along mutation carriers in established Parkinson’s risk genes, such as LRRK2 ^92,93^, iRBD has moved into the focus of biomarker development due to its high specificity for later conversion to alpha-synucleinopathies ^37^. There is significant consensus in the field that early and accurate detection of iRBD is likely the key to maximise the potential for disease modification ^30^.

Established diagnostic tools for iRBD so far involve questionnaires, which have been reported with accuracies below 70% when compared prospectively to polysomnography ^94^ and have been shown not to be valid in *de novo* Parkinson’s disease patients ^95^, or expensive and not readily available, gold-standard video-polysomnography ^70^. Until recently, only clinical, autonomic, electrophysiological and imaging biomarkers had been identified to predict pheno-conversion from iRBD to Parkinson’s disease ^37,69^. Meanwhile, CSF-based a-Syn seeding assays have been reported as one possibly accurate predictor ^25,26^. However, the invasive nature of the sampling procedure, labour-intense processing and necessary standardisation of the quantification technique is significant hindrance to a wide-spread application as a screening tool. Thus, there are currently no readily available screening tools to identify individuals at-risk of iRBD on a population level. In this context, urine proteomics appear as an ideal form of a non-invasive, easily collectable biomarker, which can be quantified in any accredited mass spectrometry lab. Sampling convenience, scalability and standardization would lend itself for the application in a population screening program.

### Summary

This study demonstrates the power of using targeted proteomics to measure multiple biomarkers at once and when combined with machine learning, a panel of biomarkers can enhance both sensitivity and specificity of tests to identify neurodegenerative disorders. This work presents a fully developed and translational targeted urine proteomic panel for the detection of iRBD. Furthermore, the assay may be run in less than 5 min/sample and can be run in any accredited, clinical mass-spectrometry laboratory and when run at scale, could cost less than $10 per sample. This panel might offer a way for future screening programs to identify individuals at high risk to develop Parkinson’s disease in very early disease stages.

**Abbreviations:** a-Syn = alpha-Synuclein; BCHE = butyrylcholinesterase; CSF = cerebrospinal fluid; ICAM1 = intercellular adhesion molecule-1; IGFBP = Insulin-like-growth-factor-binding-protein; iPD = idiopathic Parkinson’s disease; iRBD = idiopathic Rapid Eye Movement sleep behaviour disorder; ITIH2 = Inter-alpha-trypsin inhibitor heavy chain H2; LEDD = levodopa equivalent daily dose; LRRK2 (S) = symptomatic LRRK2 mutation carrier; LRRK2 (AS) = asymptomatic LRRK2 mutation carrier; MTPT = 1-Methyl-4-phenyl-1,2,3,6-tetrahydropyridin; NCAM1 = Neural cell adhesion molecule 1; NfL = neurofilament light chain; OPLS-DA = orthogonal projection to latent structures discriminant analysis; PCSK9 = Proprotein-Convertase-Subtilisin/Kexin Typ 9; PTGDS = prostaglandin D_2_ synthase; ROC = receiver operating characteristics; SPP2 = secreted phosphoprotein 2; UBC = ubiquitin C; YOPD = young onset Parkinson’s disease;

## Data Availability

All data produced in the present study are available upon reasonable request to the authors

https://panoramaweb.org/urine_irbd_trgt_proteomics.url.

## Acknowledgements

This section is not mandatory.

## Supplementary material

**Supplementary Table 1.** Demographics of samples analysed for targeted proteomics. The number of samples, percentages of males and females, and age ± standard deviation are reported for the sample groups control, iPD, iRBD, LRRK2 (S), LRRK2 (AS), YOPD and PARKIN

| Group | n | M | F | % F/M | Age ± SD |
| --- | --- | --- | --- | --- | --- |
| Controls (iPD and iRBD combined) | 66 | 35 | 31 | 47/53 | 64.7 ± 10.1 |
| Control (iPD) | 49 | 21 | 28 | 57.1/42.9 | 64.4 ± 10.6 |
| Control (iRBD) | 17 | 14 | 3 | 17.6/82.4 | 65.5 ± 9.1 |
| iPD | 59 | 35 | 24 | 40.7/59.3 | 67.1 ± 8.4 |
| iRBD | 18 | 14 | 4 | 22.2/77.8 | 67.2 ± 9.2 |
| LRRK2 (S) | 13 | 9 | 4 | 30.8/69.2 | 57.6 ± 7.5 |
| LRRK2 (AS) | 3 | 1 | 2 | 66.7/33.3 | 51.5 ± 26.2 |
| YOPD | 19 | 15 | 4 | 21.1/78.9 | 48.2 ± 7.7 |
| Parkin | 6 | 5 | 1 | 16.7/83.3 | 49.2 ± 5.0 |

iPD: idiopathic Parkinson's disease, iRBD: Rapid Eye Movement sleep disturbance disorder, LRRK2 (S): symptomatic LRRK2 mutation carrier, LRRK2 (AS): asymptomatic LRRK2 mutation carrier, YOPD: young onset Parkinson's disease, Parkin: mutations in the PRKN gene cause autosomal recessive juvenile Parkinson's disease, M: male, F: female, SD: standard deviation.

**Supplementary Table 2.**
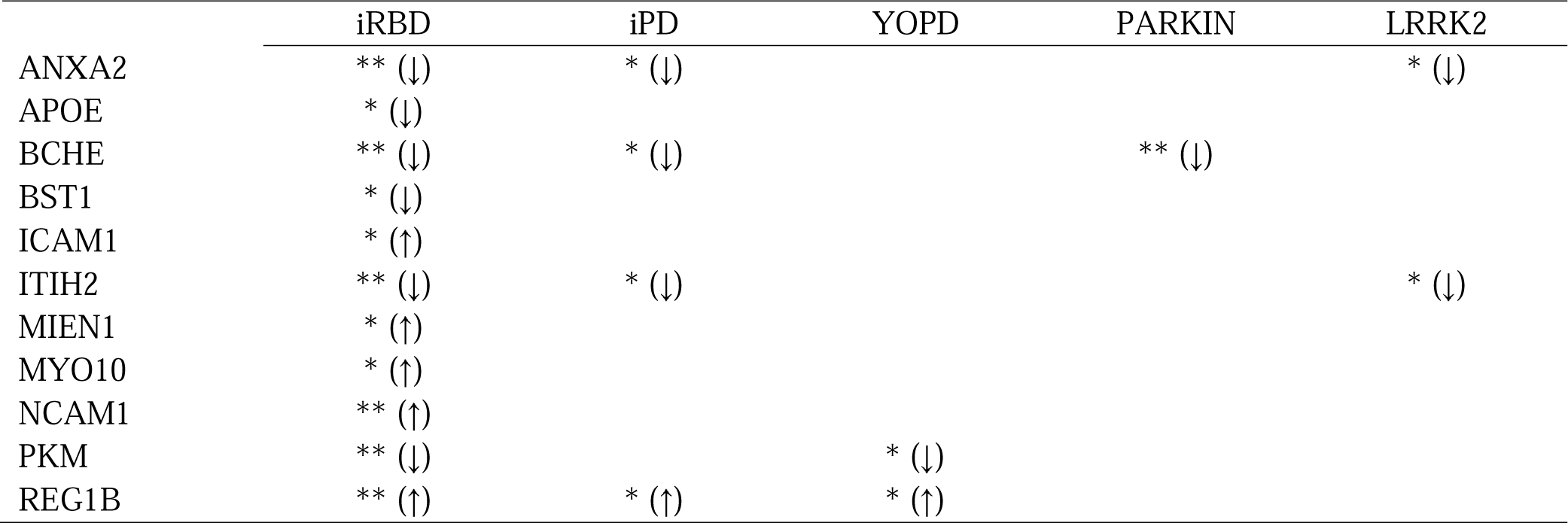
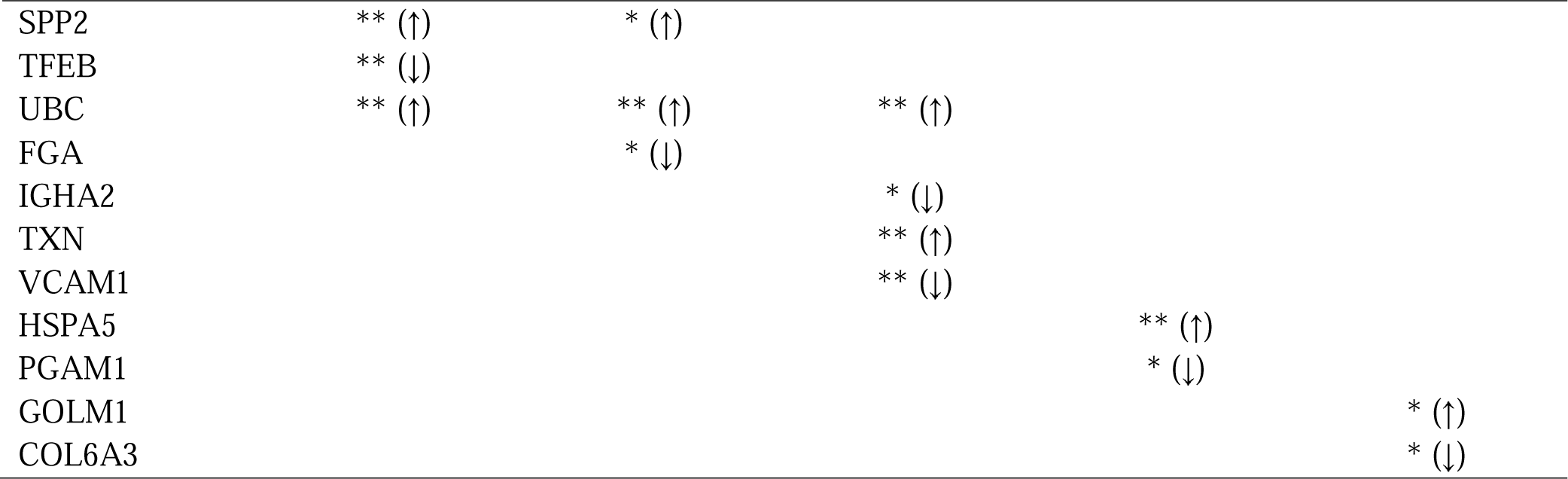
Results from the comparison between the parkinsonian groups iRBD, iPD, YOPD, PARKIN and LRRK2 *versus* control. The table shows p-value significances as ** *P* < 0.01 and * *P* < 0.05. The arrows following the asterisks represent if the protein was up- or downregulated in the disease group compared to control.

