## Supplemental Figures for "Detection of prodromal Parkinson’s disease using a urine proteomics panel and machine learning"

### Slide 1
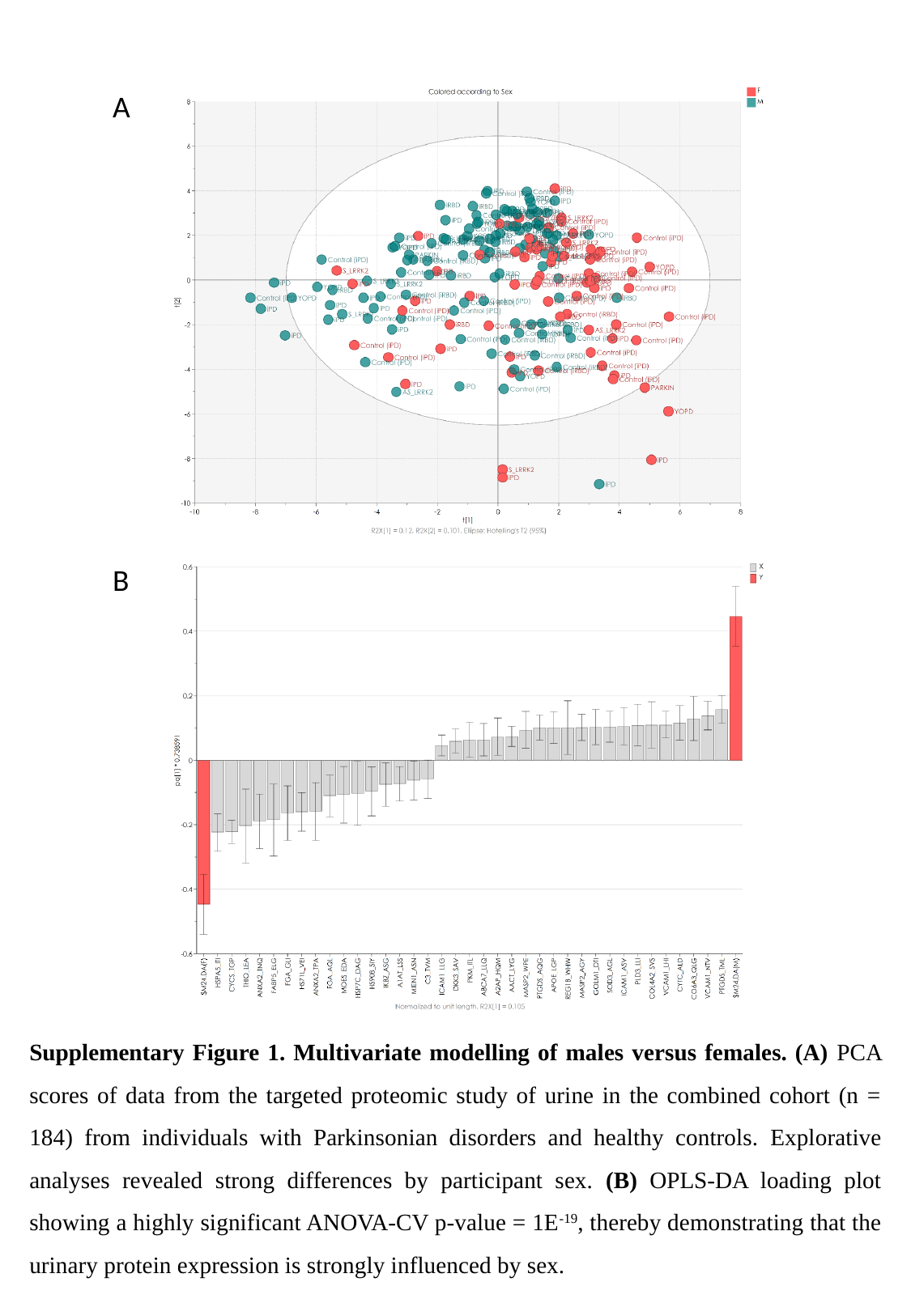

A
B
Supplementary Figure 1. Multivariate modelling of males versus females. (A) PCA scores of data from the targeted proteomic study of urine in the combined cohort (n = 184) from individuals with Parkinsonian disorders and healthy controls. Explorative analyses revealed strong differences by participant sex. (B) OPLS-DA loading plot showing a highly significant ANOVA-CV p-value = 1E^-19^, thereby demonstrating that the urinary protein expression is strongly influenced by sex.

### Slide 2
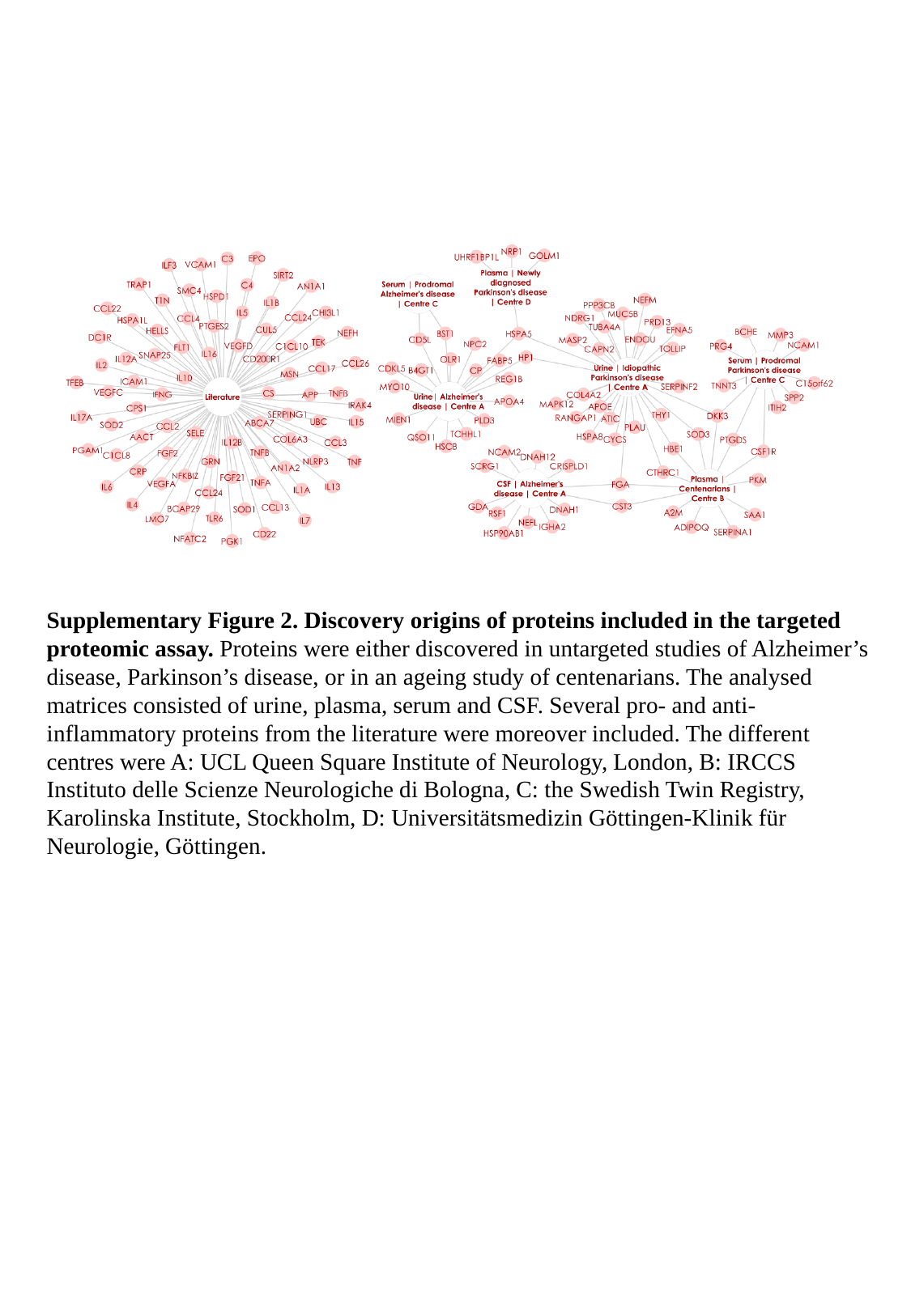

Supplementary Figure 2. Discovery origins of proteins included in the targeted proteomic assay. Proteins were either discovered in untargeted studies of Alzheimer’s disease, Parkinson’s disease, or in an ageing study of centenarians. The analysed matrices consisted of urine, plasma, serum and CSF. Several pro- and anti-inflammatory proteins from the literature were moreover included. The different centres were A: UCL Queen Square Institute of Neurology, London, B: IRCCS Instituto delle Scienze Neurologiche di Bologna, C: the Swedish Twin Registry, Karolinska Institute, Stockholm, D: Universitätsmedizin Göttingen-Klinik für Neurologie, Göttingen.

### Slide 3
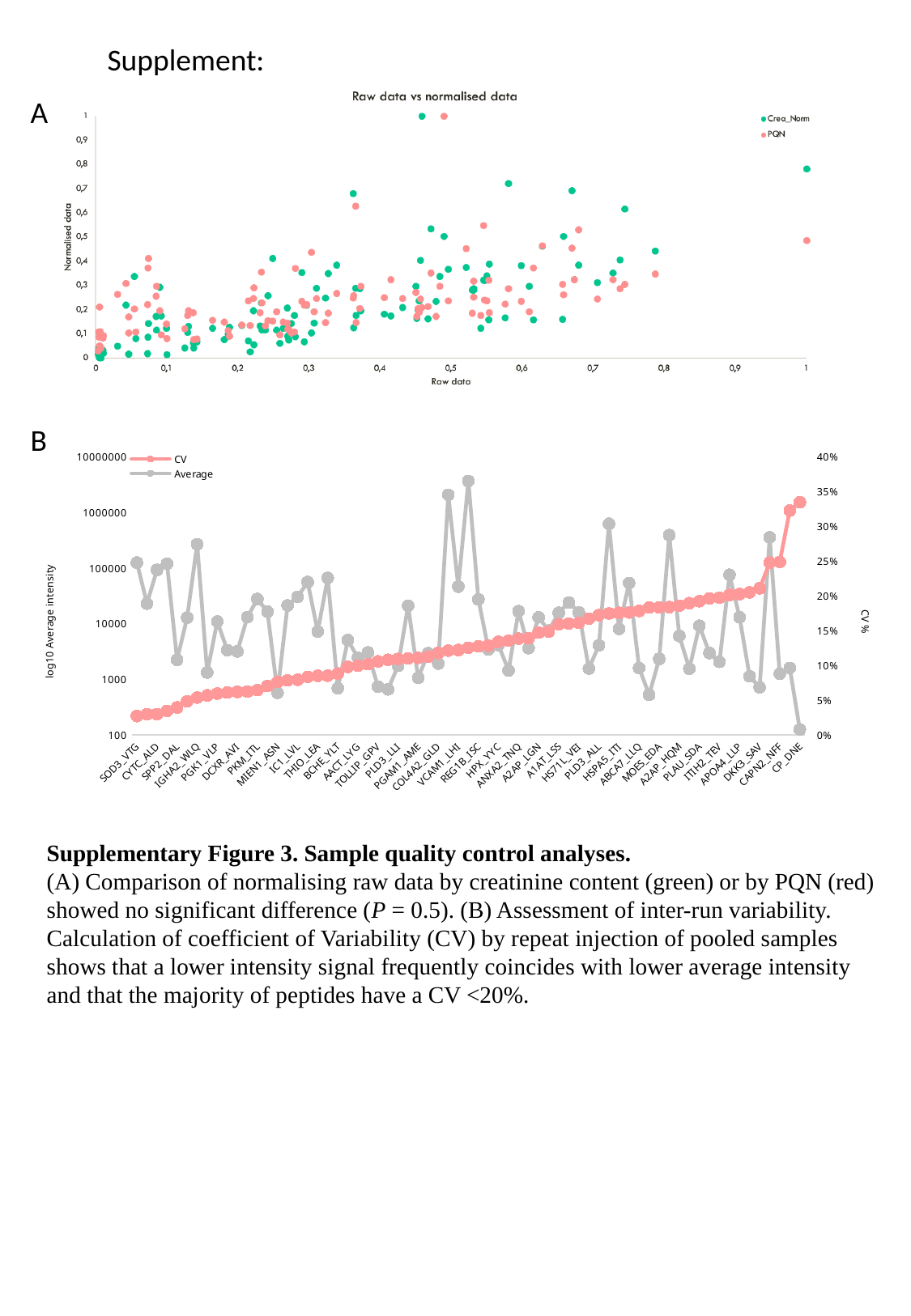

Supplement:
A
B
#### Chart
| Category | Average | CV |
|---|---|---|
| SOD3_VTG | 124138.50954755559 | 0.026801009789260196 |
| FGA_AQL | 22773.921325466992 | 0.029482153001022138 |
| CYTC_ALD | 91693.30642154823 | 0.029635453588499977 |
| REG1B_WHW | 118453.28076909836 | 0.03414784984743577 |
| SPP2_DAL | 2215.2047641971462 | 0.03913650434777904 |
| HSP7C_DAG | 12707.5074168489 | 0.04805146390064566 |
| IGHA2_WLQ | 265785.54966519563 | 0.053488151586328 |
| ITIH2_VQF | 1311.384751646601 | 0.056530558940397345 |
| PGK1_VLP | 10800.875694947528 | 0.059199447622045896 |
| TOLLIP_LNI | 3300.4950604725323 | 0.06058988182527696 |
| DCXR_AVI | 3138.8239505392075 | 0.06146557241526882 |
| COL4A2_SVS | 12981.395575677148 | 0.061926352442917845 |
| PKM_ITL | 27436.893899362243 | 0.06406860522380453 |
| APOA4_LAP | 16271.148610472574 | 0.0700305129872733 |
| MIEN1_ASN | 565.1390962694405 | 0.07580662820446317 |
| ICAM1_LLG | 21103.997085118648 | 0.07804365293873745 |
| IC1_LVL | 30025.697033430086 | 0.07908847024390424 |
| VCAM1_NTV | 55416.54107793321 | 0.08314916348607343 |
| THIO_LEA | 7140.196091902213 | 0.08470222109167644 |
| GOLM1_DTI | 65779.29286870164 | 0.08490557753361565 |
| BCHE_YLT | 684.9883181641743 | 0.08792482784740815 |
| CYCS_TGP | 5017.97950954011 | 0.0973604176268271 |
| AACT_LYG | 2407.4659790415217 | 0.09923908103901476 |
| CO6A3_QLG | 3008.2647909885604 | 0.10138735292251687 |
| TOLLIP_GPV | 729.8047514681517 | 0.10536945047973964 |
| C3_TVM | 652.5974935409474 | 0.10761713848320895 |
| PLD3_LLI | 1730.489869324572 | 0.10891737673791707 |
| UBC_TIT | 20824.946253260692 | 0.10981882620661039 |
| PGAM1_AME | 1054.2417690351795 | 0.11043709357626148 |
| FGA_GLI | 2913.2618354316655 | 0.11229496510222804 |
| COL4A2_GLD | 1899.8634519540426 | 0.11737999067131107 |
| PTGDS_AQG | 2050161.4124903008 | 0.12090021378430472 |
| VCAM1_LHI | 45893.65026606625 | 0.1218660034062778 |
| PTGDS_TML | 3620834.2582363826 | 0.12497620778474008 |
| REG1B_ISC | 27039.208866887213 | 0.12713248856775006 |
| ICAM1_ASV | 3398.7324966217393 | 0.12795127621578967 |
| HPX_YYC | 4078.2113373145316 | 0.13343659982593534 |
| QSOX1_LEE | 1427.1119360209786 | 0.13505380205202414 |
| ANXA2_TNQ | 16564.664104943935 | 0.138058026228658 |
| B4GT1_YWL | 3590.7944956076594 | 0.13874449745475984 |
| A2AP_LGN | 12856.527294595475 | 0.14673765438911526 |
| ANXA2_TPA | 7435.172978049303 | 0.1483558781764496 |
| A1AT_LSS | 15566.518574057245 | 0.15891545117820116 |
| MIEN1_EQY | 23810.486867237047 | 0.15994503364703258 |
| HS71L_VEI | 15807.26549140551 | 0.16080341958189623 |
| APOE_LGA_E4 | 1545.4399240897553 | 0.16687498609372325 |
| PLD3_ALL | 4036.5036543973056 | 0.17192243770161833 |
| FABP5_ELG | 617253.7330423415 | 0.17436818940968163 |
| HSPA5_ITI | 8001.249837988781 | 0.17522882024349995 |
| SOD3_AGL | 53181.31441295138 | 0.17577395080678304 |
| ABCA7_LLQ | 1576.9691247754986 | 0.17799677543520734 |
| APP_GLT | 522.3553937355783 | 0.18283924343477168 |
| MOES_EDA | 2311.791466222056 | 0.18302780664457297 |
| MASP2_WPE | 385311.2442382509 | 0.18343331203470453 |
| A2AP_HQM | 5938.445716658874 | 0.18556437533138867 |
| IKBZ_ASG | 1535.1950663011075 | 0.1891397552615084 |
| PLAU_SDA | 9073.267133943264 | 0.19203653768221843 |
| CH60_VTD | 2937.324688857759 | 0.19595687486776392 |
| ITIH2_TEV | 2041.3342436386215 | 0.19706393324809893 |
| APOE_LGP | 74796.57439963041 | 0.20088127624384836 |
| APOA4_LLP | 12929.58349335192 | 0.20233500872131444 |
| NCAM1_LEG | 1123.8628148788348 | 0.2047248884898371 |
| DKK3_SAV | 709.2577314113041 | 0.21048353593223126 |
| MASP2_AGY | 351932.69141936634 | 0.2473799469694761 |
| CAPN2_NFF | 1242.8984509142201 | 0.24841176590259337 |
| HS90B_SIY | 1576.686501951609 | 0.32228736608197306 |
| CP_DNE | 124.57470491911612 | 0.33413205315771344 |Supplementary Figure 3. Sample quality control analyses.
(A) Comparison of normalising raw data by creatinine content (green) or by PQN (red) showed no significant difference (P = 0.5). (B) Assessment of inter-run variability. Calculation of coefficient of Variability (CV) by repeat injection of pooled samples shows that a lower intensity signal frequently coincides with lower average intensity and that the majority of peptides have a CV <20%.

### Slide 4
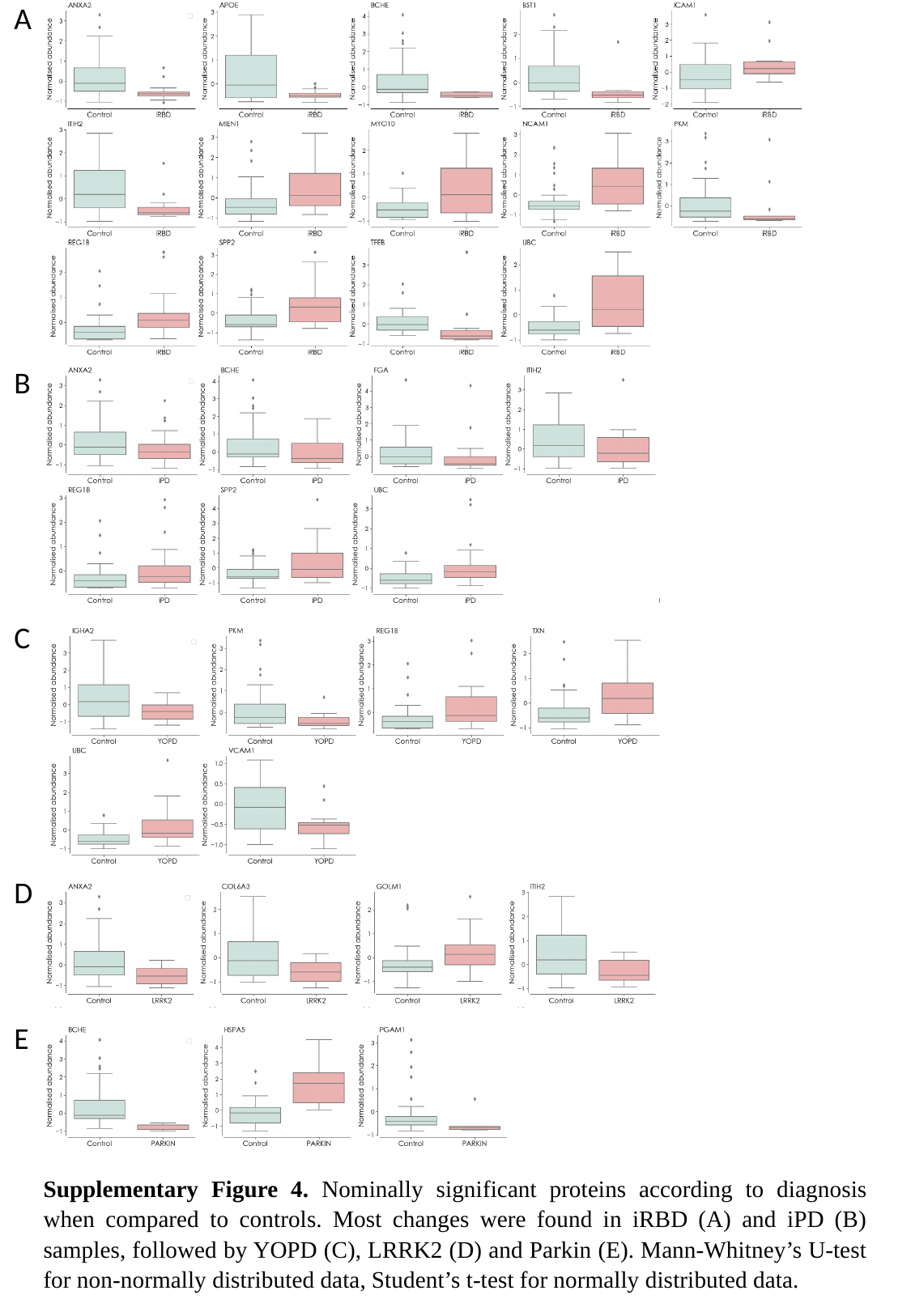

A
B
C
D
E
Supplementary Figure 4. Nominally significant proteins according to diagnosis when compared to controls. Most changes were found in iRBD (A) and iPD (B) samples, followed by YOPD (C), LRRK2 (D) and Parkin (E). Mann-Whitney’s U-test for non-normally distributed data, Student’s t-test for normally distributed data.

### Slide 5
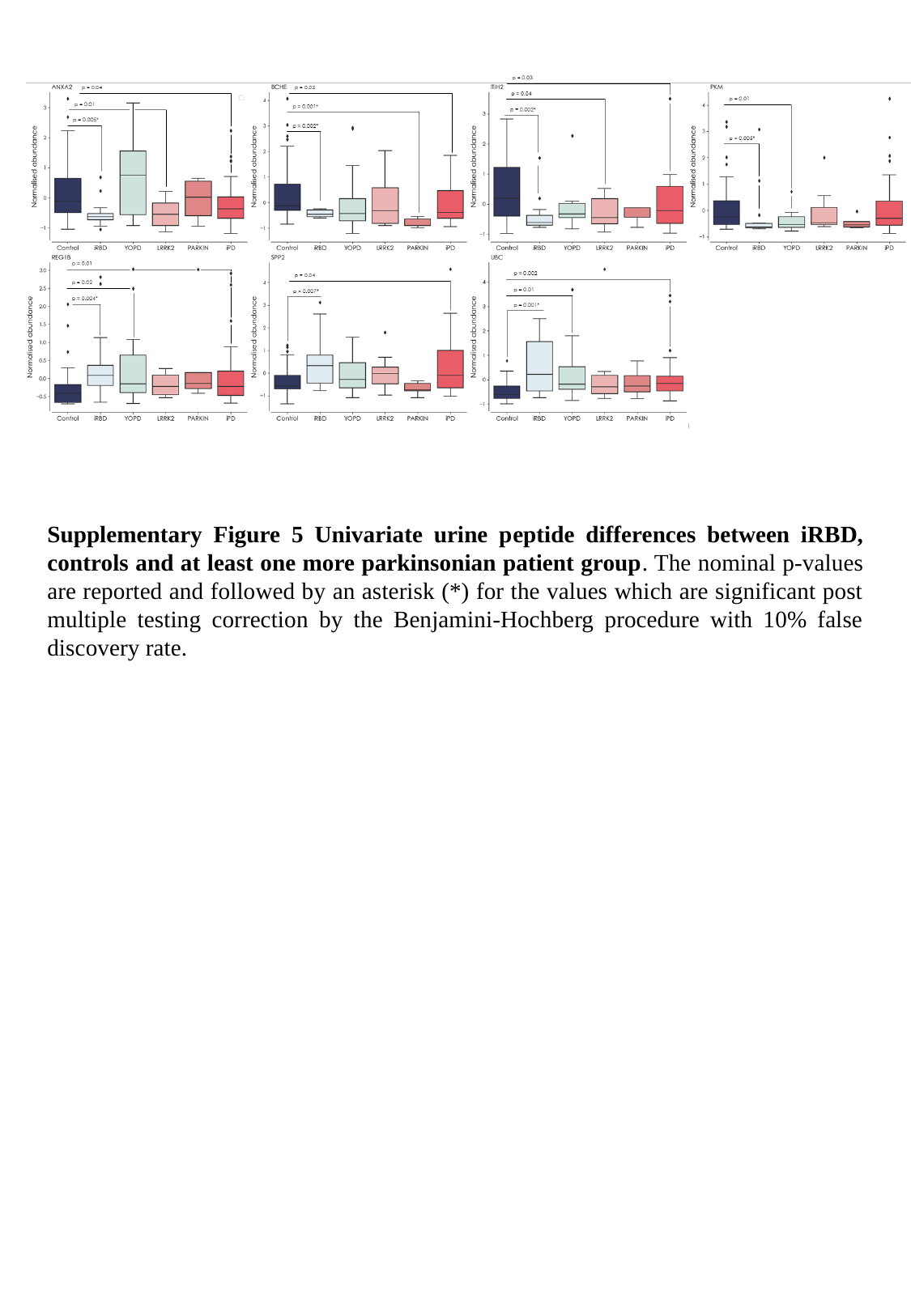

Supplementary Figure 5 Univariate urine peptide differences between iRBD, controls and at least one more parkinsonian patient group. The nominal p-values are reported and followed by an asterisk (*) for the values which are significant post multiple testing correction by the Benjamini-Hochberg procedure with 10% false discovery rate.

### Slide 6
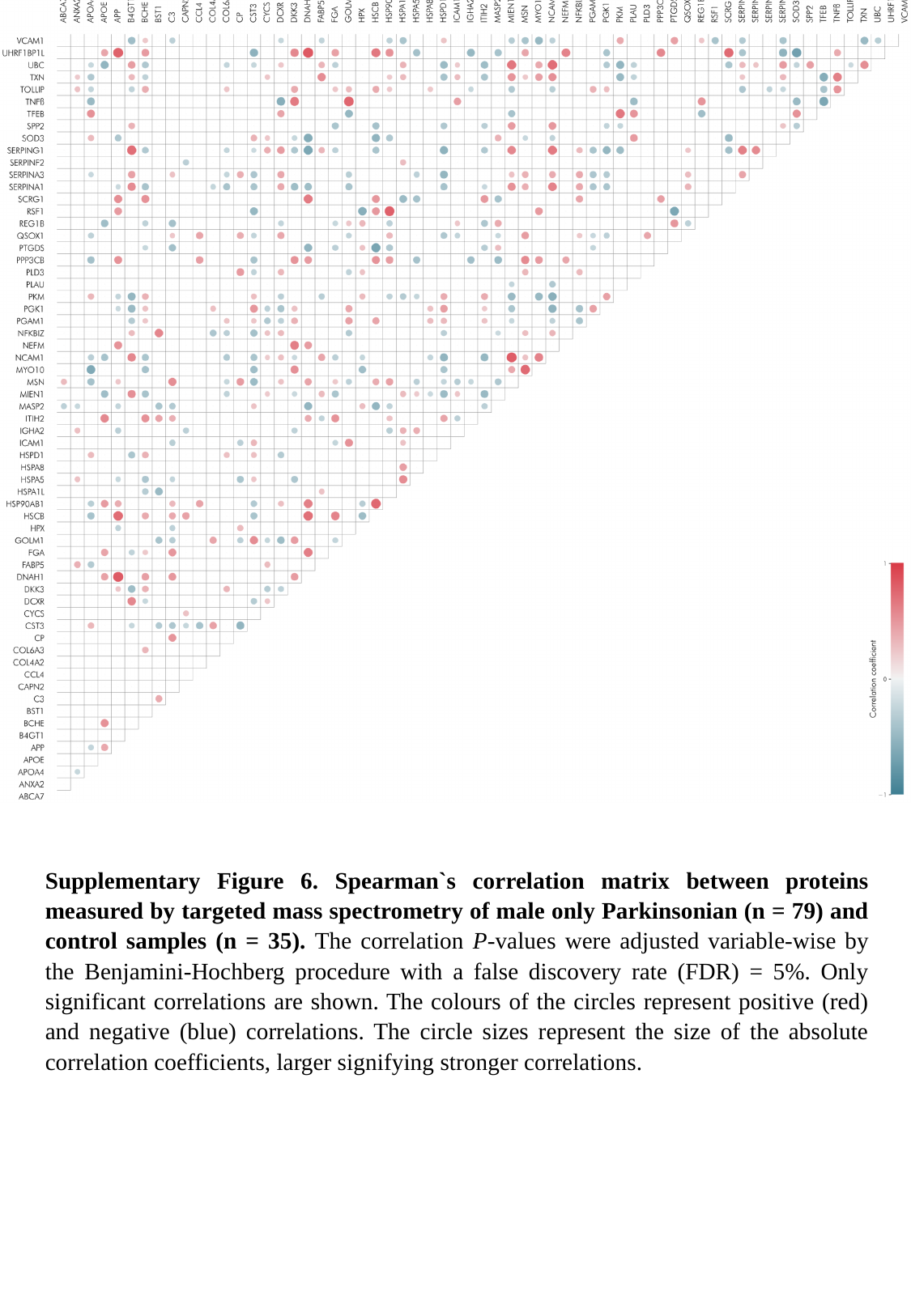

Supplementary Figure 6. Spearman`s correlation matrix between proteins measured by targeted mass spectrometry of male only Parkinsonian (n = 79) and control samples (n = 35). The correlation P-values were adjusted variable-wise by the Benjamini-Hochberg procedure with a false discovery rate (FDR) = 5%. Only significant correlations are shown. The colours of the circles represent positive (red) and negative (blue) correlations. The circle sizes represent the size of the absolute correlation coefficients, larger signifying stronger correlations.
